# One year of modeling and forecasting COVID-19 transmission to support policymakers in Connecticut

**DOI:** 10.1101/2020.06.12.20126391

**Authors:** Olga Morozova, Zehang Richard Li, Forrest W. Crawford

**Affiliations:** Program in Public Health and Department of Family, Population and Preventive Medicine, Stony Brook University (SUNY), NY, USA; Department of Statisitcs, University of California, Santa Cruz, Santa Cruz, CA, USA; Department of Biostatistics, Yale School of Public Health, New Haven, CT, USA; Department of Statistics & Data Science, Yale University, New Haven, CT, USA; Department of Ecology & Evolutionary Biology, Yale University, New Haven, CT, USA; Yale School of Management, New Haven, CT, USA

**Keywords:** SEIR epidemic model, SARS-CoV-2, social distancing, effective reproduction number, case detection ratio, infection fatality ratio, infection hospitalization ratio

## Abstract

To support public health policymakers in Connecticut, we developed a county-structured compartmental SEIR-type model of SARS-CoV-2 transmission and COVID-19 disease progression. Our goals were to provide projections of infections, hospitalizations, and deaths, as well as estimates of important features of disease transmission, public behavior, healthcare response, and clinical progression of disease. In this paper, we describe a transmission model developed to meet the changing requirements of public health policymakers and officials in Connecticut from March 2020 to February 2021. We outline the model design, implementation and calibration, and describe how projections and estimates were used to support decision-making in Connecticut throughout the first year of the pandemic. We calibrated this model to data on deaths and hospitalizations, developed a novel measure of close interpersonal contact frequency to capture changes in transmission risk over time and used multiple local data sources to infer dynamics of time-varying model inputs. Estimated time-varying epidemiologic features of the COVID-19 epidemic in Connecticut include the effective reproduction number, cumulative incidence of infection, infection hospitalization and fatality ratios, and the case detection ratio. We describe methodology for producing projections of epidemic evolution under uncertain future scenarios, as well as analytical tools for estimating epidemic features that are difficult to measure directly, such as cumulative incidence and the effects of non-pharmaceutical interventions. The approach takes advantage of our unique access to Connecticut public health surveillance and hospital data and our direct connection to state officials and policymakers. We conclude with a discussion of the limitations inherent in predicting uncertain epidemic trajectories and lessons learned from one year of providing COVID-19 projections in Connecticut.

## Introduction

Epidemiologic models of infectious disease transmission have played an important role in supporting public health decision-making during the COVID-19 pandemic [1–7]. By specifying structural features of infection transmission dynamics, models can provide insights into epidemiologic parameters, historical trends in epidemic dynamics, or future outcomes under hypothetical intervention scenarios. Transmission models are especially useful in situations of high uncertainty, offering a structured way to assess the potential effects of interventions given plausible assumptions about disease transmission. Transmission models may also be useful for short-term forecasting: when it is feasible to assume that key epidemiologic features will remain constant over time, such models can provide projections of natural transmission dynamics given the current state of an epidemic. Models cannot predict the future with certainty, but they can be helpful for scenario analysis by bounding the range of plausible future trajectories [8]. At the same time, simple models may have poor inferential and predictive performance if they fail to capture important features of disease transmission that may vary over time.

In early 2020, many countries, including the US, faced a public health crisis caused by the COVID-19 pandemic. In places like New York City and large cities in California, COVID-19 cases increased rapidly. In New York City, severely ill patients overwhelmed hospitals [9, 10] with death rates as high as 9% among confirmed cases and 32% among hospitalized patients [11]. Policymakers from the US regions first affected by the pandemic, Connecticut being one of them, were unprepared for its magnitude and severity. In the absence of effective pharmaceutical interventions, state and local governments turned to public health control measures that were last widely used during the 1918 influenza pandemic, such as social distancing and stay-at-home orders, to slow transmission of SARS-CoV-2. As transmission subsided, states began considering phased lifting of social distancing restrictions. Several urgent questions emerged: 1) How soon can interventions like school closures and stay-at-home orders be lifted? 2) How should public health interventions be implemented to minimize the risk of a resurgence? 3) What will be the effect of phased reopening plans on cases, hospitalizations, and deaths? Surveillance data on testing, case counts, hospitalizations, and deaths was useful in characterizing the dynamics of the initial wave, but policymakers needed predictive analytic tools to evaluate the current state of an epidemic and asess the risk of future resurgence.

A wide variety of transmission models were developed during the early months of the COVID-19 pandemic. These models were constructed for several related purposes. Many models sought to estimate basic epidemiologic parameters including basic reproduction number (*R*_0_), epidemic growth rate and doubling time, serial interval, case and infection fatality ratios, and case detection ratio [6, 12–20]. Some models produced simulated future outcomes under hypothetical behavioral or interventional scenarios [1, 4, 21]. Other models had a retrospective inferential goal of estimating effects of past interventions including lockdown and other non-pharmaceutical measures [6, 14, 16, 18, 20]. Some early models did not calibrate parameters to data, and instead simulated from models parameterized using published estimates from early observational studies or other infections with similar properties [1, 4, 21]. Intervention effects were often included as constant model parameters or estimated by calibrating the model separately to pre- and post-intervention time periods [6, 15, 16]. In the early stages of the pandemic, these models helped demonstrate the dangers of unmitigated transmission and provided some evidence of the effectiveness of non-pharmaceutical interventions. However most of these models relied on publicly available data, often limited to official case counts, from the first wave of an epidemic [12–20], and assumed constant transition rates (other than reduction in transmission following initial lockdown), and were therefore unable to adequately capture changing features of the COVID-19 burden, including behavioral changes, time-varying policy response, clinical management of the disease, or healthcare system dynamics [1, 13–16, 19, 21].

As the initial epidemic wave in Connecticut began to subside during the summer of 2020, local policymakers needed models that could answer specific questions about past and current infection dynamics, accommodate established epidemiologic features of disease transmission, permit prediction of outcomes under policy scenarios identified by stakeholders, and provide projections of policy-relevant outcomes under assumptions that could be understood by policymakers. Several nationwide forecasting tools were developed to provide state- or county-level projections, relying on data universally and publicly available across all locations [3, 7, 22]. These models employed assumptions applied universally across all locations and provided a limited set of outputs that were not always able to address the needs of local policymakers. These factors motivated the development of transmission models tailored to local context, the timing of intervention events and planned future policy changes, and data that were only available at the local level [23–26].

In this paper, we present a county-structured model of SARS-CoV-2 transmission and COVID-19 disease progression in Connecticut. The model was developed and improved over the first year of the pandemic to support decision-making by Connecticut public health officials and policymakers [27]. This work is the result of continuous feedback from Connecticut public health officials, who provided detailed data on congregate and non-congregate testing, cases, and deaths, age-stratified cases, and hospitalization admissions and census – a feature that is absent from all nationwide analyses. We first describe the epidemic and public policy response in Connecticut, emphasizing the inferential questions articulated by public officials during development of the model. We then briefly describe the structure of the transmission model, its parameters, and data sources used for model calibration, with full details given in the Supplementary Material. We present results, including model predictions of hospitalizations and deaths on the dates of important decisions made by policymakers, estimates of the number of COVID-19 infections, case-detection rate, cumulative incidence, effective reproduction number, infection fatality ratio, and other epidemiologic parameters. We conclude with a discussion of the limitations inherent in predicting uncertain epidemic trajectories using models, and outline lessons learned from one year of providing COVID-19 projections to support Connecticut policymakers.

### The COVID-19 epidemic and response in Connecticut

Connecticut (population 3.565 million) was among the US states most severely impacted by the first wave of COVID-19 epidemic [28]. On March 8th, 2020, the first Connecticut COVID-19 case was reported, followed by a rapid increase in case counts. In the first three weeks of the epidemic, the state reported over 2,500 confirmed COVID-19 cases [29]. A similar rate of increase in hospitalizations followed, and on April 2, 2020, COVID-19 hospitalization census exceeded 1,000. On March 17, Governor Ned Lamont ordered all in-person classes at K-12 schools canceled, and later extended the closure for the remainder of the 2019-2020 academic year [30–33]. The Governor issued a statewide “Stay Safe, Stay Home” order to take effect on March 23 [34]. The order called on all nonessential businesses to cease in-person operations. Essential businesses could remain open with additional restrictions and guidelines to minimize close contact and risk of transmission. Evidence from mobile device data suggests that Connecticut residents reduced their mobility before the official lockdown order went in effect [35].

The number of hospitalized COVID-19 patients in Connecticut peaked on April 21, 2020 and began a slow decline [29]. In early May, Governor Lamont issued plans and guidance for reopening, a process set to begin with “Phase 1” on May 20 when some businesses, mostly those operating outdoors, were allowed to reopen at 50% capacity [36]. Around the same time, we released a report that included COVID-19 transmission projections through August 31, 2020 under different scenarios of potential contact increase during the summer [27]. Phase 2 of reopening began on June 17th when indoor dining, libraries and religious services were allowed to reopen at reduced capacity [37]. Reopening was followed by scale-up in testing: average daily number of polymerase chain reaction (PCR) tests increased from about 2,000 at the beginning of April to about 8,000 at the end of June [29].

For most of summer 2020, case counts and hospitalizations in Connecticut remained low, even while large outbreaks were happening in many parts of the US [29]. The major state-level policy question during this time was whether and how to reopen primary, secondary, and college/university schooling in the fall. At the end of August, we developed model projections of infections, hospitalizations, and deaths for fall under different assumptions about the rate of close interpersonal contact associated with reopening of schools. These forecasts predicted increasing infections and a statewide resurgence during the fall 2020. In-person, remote, and hybrid education at all levels resumed in Connecticut in August and early September. Case counts and hospitalizations during fall 2020 increased slowly [29], leading the Governor to implement Phase 3 reopening, permitting indoor businesses to operate at higher capacity, on October 8 [38]. By early November, public health officials recognized a broad statewide epidemic resurgence. In response to rising case counts and fears of a substantial second wave, on November 6 Governor Lamont reverted to “Phase 2.1”, reducing permitted occupancy of indoor businesses and events [39]. In-person education at most public schools and all universities ended in mid-November before the Thanksgiving recess. Asymptomatic testing programs implemented by many universities were subsequently scaled down, resulting in a reduction in testing rates in Connecticut – making it difficult to interpret changes in the test positive proportion. Case counts in the second wave of the epidemic peaked in mid-December, plateaued for about a month and began a slow decline starting the second half of January 2021.

As of mid-March 2021, most schools and universities have reopened with a simultaneous substantial increase in close contact rates. Vaccine deployment in Connecticut began on December 14th, 2020 with residents of congregate settings being vaccinated first. As of March 1, 2021, 8.9% of Connecticut population received full vaccination schedule and 16.5% received at least one dose of the vaccine with most vaccines being administered among residents of congregate settings and in the age group of 75 year old and above [29].

## Methods

### Data sources

Our modeling approach relies on multiple data streams provided by the Connecticut Department of Public Health (CT DPH) and the Connecticut Hospital Association (CHA). Some of these datasets are publicly available, while others, including public health surveillance data, were obtained through a contract agreement between CT DPH and the Yale School of Public Health. Baseline non-institutionalized county-level populations and age demographics in Connecticut were obtained from the American Community Survey [40]. Figure 1 shows data series, described below, used in model parametrization and calibration.

**Figure 1:**
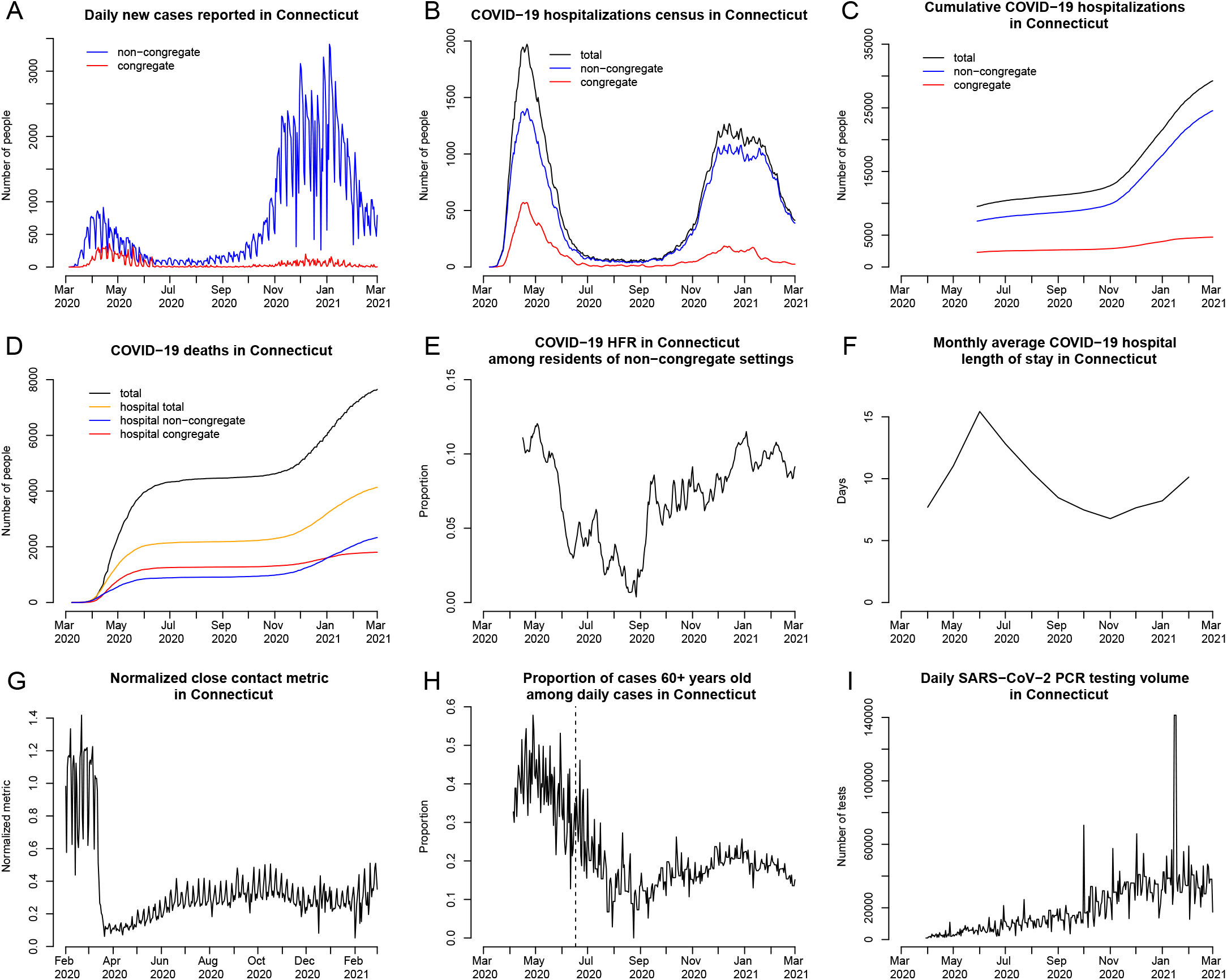
Observed and estimated data used in model calibration and approximation of time-varying model parameters. Top row: A: daily new cases reported in Connecticut by the date of specimen collection among residents of non-congregate and congregate settings; B: COVID-19 hospitalization census; C: cumulative COVID-19 hospitalizations. In plots B and C, the total number (black line) represents observed data, while non-congregate (blue) and congregate (red) lines represent estimates. Middle row: D: cumulative COVID-19 deaths in Connecticut; E: hospital case fatality ratio (HFR) among hospitalized residents of non-congregate settings (estimated); F: average length of hospital stay among COVID-19 patients by month. Bottom row: G: normalized close interpersonal contact metric relative to the pre-epidemic period; H: proportion of cases 60+ years old among daily COVID-19 cases (only data on the right of the dashed line is used in model parametrization); I: daily PCR testing volume.

### Hospitalization, deaths, and hospital capacity

We obtained data on daily confirmed COVID-19 hospitalization census, cumulative COVID-19 hospitalizations, cumulative number of deaths among hospitalized patients, and daily total available hospital beds (including occupied) in Connecticut from CHA [41]. Hospitalizations census and deaths time series are available since the epidemic onset in March 2020. CHA changed their data collection and reporting procedures with respect to admissions and discharges in late May 2020, and cumulative hospitalizations data are available starting May 29, 2020. Data on the total number of COVID-19 deaths are publicly available and were obtained from the Connecticut Open Data Portal [29].

The transmission model aims to capture community spread of SARS-CoV-2, and therefore excludes transmission occurring in congregate settings like skilled nursing and assisted living facilities, or prisons. Similar to Salje et al. [6], we excluded congregate settings, since transmission in small closed communities violates important modeling assumptions related to mixing patterns in the population. Available hospitalization data do not disaggregate by the patient’s place of residence at the time of diagnosis or hospitalization. According to CT DPH, as of October 30, 2020, about 73% of all deaths have occurred among residents of congregate settings, primarily nursing homes, emphasizing that the first wave of the epidemic was heavily dominated by transmission in this population. To address this issue, we estimated the time series of hospitalizations (census and cumulative) and hospital deaths coming from non-congregate settings and used these estimated counts in the model calibration. We received data on daily COVID-19 death counts in hospitals disaggregated by the type of residence (congregate vs. non-congregate) at the time of diagnosis or hospitalization from CT DPH (Plot D in Figure 1). Based on these data, we estimated the time-varying proportion of hospitalization census and cumulative hospitalizations coming from congregate and non-congregate settings. A detailed description of this process is provided in the Supplement. Plots B and C in Figure 1 show estimates of these time series along with observed total numbers.

Data on estimated daily hospital admissions and deaths from non-congregate settings were used to estimate time-varying hospital case fatality ratio (HFR) and used as a time-varying parameter in the transmission model (Plot E in Figure 1). Data on monthly average hospital length of stay among COVID-19 inpatients in Connecticut hospitals were provided by the CHA and used as a time-varying parameter in the transmission model (Plot F in Figure 1).

### COVID-19 tests, cases, and age distribution of cases

We assume that widespread testing shortens the time between infection and diagnosis and may therefore lead to shorter duration of transmissibility via isolation of infected individuals. We use daily PCR testing volume (Plot I in Figure 1) to parameterize the time-varying duration of infectiousness among mildly symptomatic and asymptomatic cases. Data on daily PCR tests were obtained from the Connecticut Open Data Portal [29].

We used the proportion of daily confirmed COVID-19 cases aged 60 years old and above to parameterize the dynamics of severe infections over time (Plot H in Figure 1). Early in the epidemic, testing was not widely available and was primarily used to confirm severe cases that were more likely to be among older people. Therefore, in model parametrization, we assume a constant severe proportion early in the epidemic, and use these data to approximate changing severity proportion beyond the second phase of reopening, which started on June 17, 2020, when testing became widely available (vertical dashed line in Plot H in Figure 1). Plot A in Figure 1 shows reported case counts in Connecticut by residence type (congregate or non-congregate).

### Close interpersonal contact

Close interpersonal contact (within six feet) is the main route for transmission of SARS-CoV-2 [42]. Social distancing interventions implemented in Connecticut were intended to reduce the frequency of such contact. We therefore estimated the frequency of close interpersonal contact everywhere in Connecticut using mobile device geolocation data. The project, described separately by Crawford et al. [43], developed a novel probabilistic measure of contact and aggregated contact events at the town and state levels to describe the dynamics of close interpersonal contact. Plot G in Figure 1 shows that statewide close contact in Connecticut dropped from its February 2020 baseline about one week prior to the Governor’s stay-at-home order, and rose slowly throughout the summer and fall.

### Compartmental model

We developed a deterministic compartmental model of SARS-CoV-2 transmission and COVID-19 disease progression. The model is based on the SEIR (susceptible, exposed, infectious, removed) framework [44], which we extend to accommodate geographical variation in Connecticut, hospital capacity, and distinctive features of COVID-19 disease. The model is implemented at the level of individual counties in Connecticut and assumes that most transmission occurs within a given county. A small proportion of contacts (1.5%) is allowed to happen between adjacent counties.

Figure 2 shows a schematic representation of the transmission model structure within a single county, the county map of Connecticut [45], and the county adjacency matrix.

**Figure 2:**
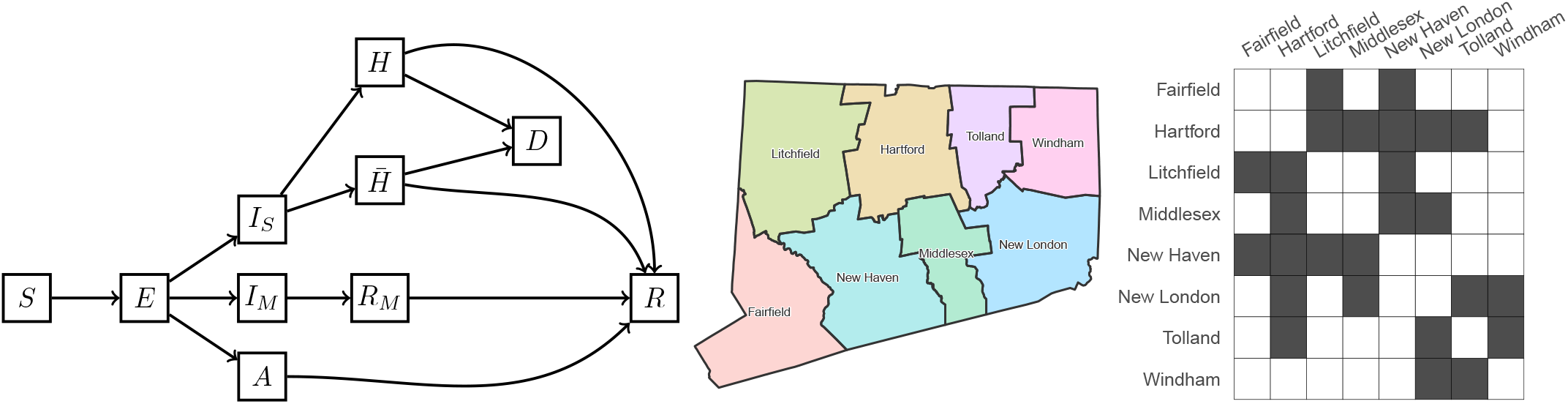
Schematic illustration of the model of SARS-CoV-2 transmission and COVID-19 disease progression; county map of Connecticut and county adjacency matrix. Individuals begin in the susceptible (*S*) compartment. Exposed individuals (*E*) may develop either asymptomatic (*A*), mild (*I*_*M*_), or severe (*I*_*S*_) infection. Asymptomatic and mild infections resolve without hospitalization and do not lead to death. Mild symptomatic cases self-isolate (*R*_*M*_) shortly after development of symptoms, and transition to recovery (*R*) when infectiousness ceases. All severe cases require hospitalization (*H*) unless hospitalization capacity is exhausted, in which case they transition to 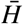 representing hospital overflow, then to recovery (*R*) or death (*D*). The model captures infection transmission in non-congregate settings, and excludes cases and deaths occurring in settings like nursing homes and prisons. It assumes a closed population without births and does not capture non-COVID-19 deaths. In the adjacency matrix, the dark gray cells correspond to counties that are adjacent.

We categorize infections as asymptomatic (*A*), mild symptomatic (*I*_*M*_), and severe (*I*_*S*_). Only severe infections may lead to death (*D*). Severe infections are defined as those requiring hospitalization (*H*). If hospitalization capacity is overwhelmed, severe cases in the community are denied hospitalization 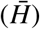, and experience a higher probability of death compared to hospitalized cases. Mild symptomatic cases are assumed to self-isolate shortly after they develop symptoms (*R*_*M*_) and remain isolated until they recover (*R*). The average time that severe cases spend in the infectious state is approximated by the time between onset of infectiousness and hospitalization (or attempted hospitalization in case of hospital overflow). The force of infection from hospitalized patients to unhospitalized susceptible individuals is assumed to be negligible. We further assume that recovered individuals remain immune to reinfection for the duration of the study period. Let *N*_*i*_ be the population size of county *i* and let *J*_*i*_ be the set of counties adjacent to county *i*. Let *C* ^(*i*)^ represent hospitalization capacity in county *i*, which may vary over time. Transmission dynamics in county *i* are given by the following system of ordinary differential equations:

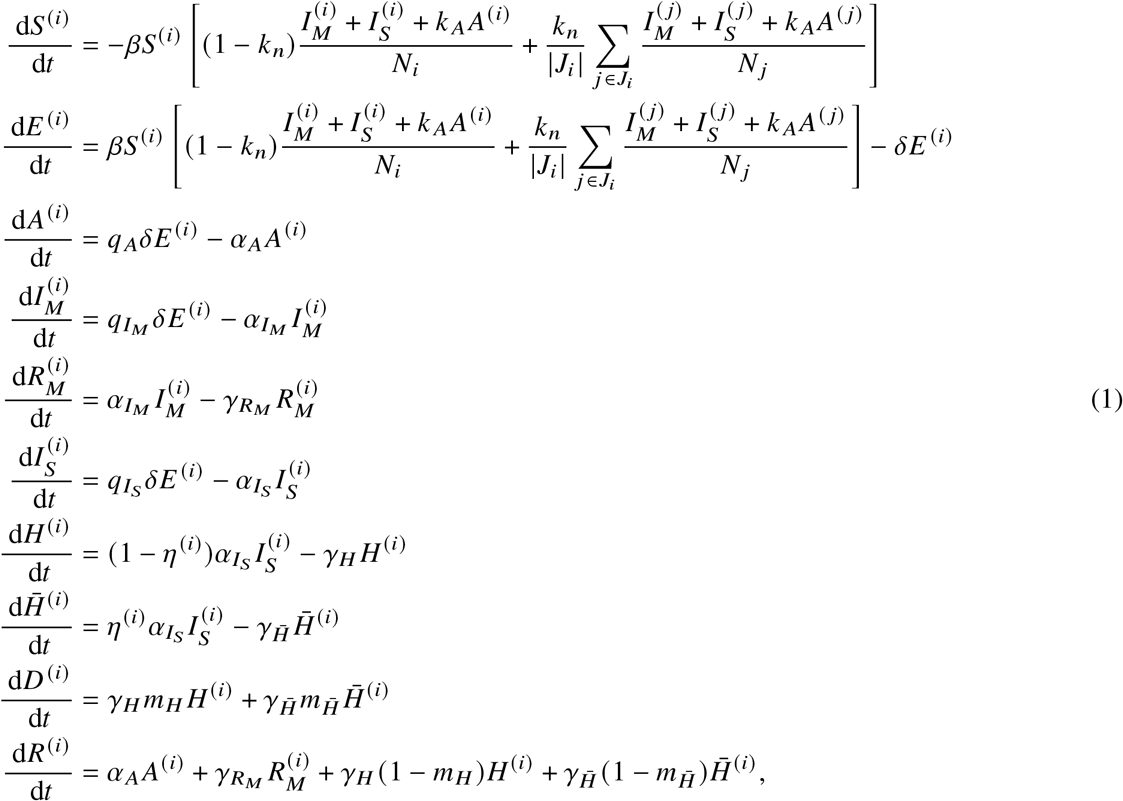

where 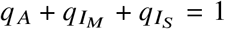. The function *η*^(*i*)^ = [1 + exp(0.5(*C*^(*i*)^ −*H*^(*i*)^))]^−1^ is a “soft” hospitalization capacity overflow function. Table 1 lists model parameters and their definitions. The analysis was performed using the R statistical computing environment [46, 47].

**Table 1:** Transmission model parameters.

| Notation | Definition |
| --- | --- |
| $\beta$ | Transmission parameter per susceptible - infectious pair |
| $\delta$ | 1 / Latency period (days<sup>-1</sup>) |
| $q_A, q_{I_M}, q_{I_S}$ | Proportions of infections that are asymptomatic, mild symptomatic, and severe, $q_A + q_{I_M} + q_{I_S} = 1$ |
| $\alpha_A$ | 1 / Duration of infectiousness among asymptomatic cases (days<sup>-1</sup>) |
| $k_A$ | Relative infectiousness of asymptomatic cases compared to symptomatic |
| $\alpha_{I_M}$ | 1 / Duration of infectiousness among mild symptomatic cases, time until isolation (days<sup>-1</sup>) |
| $\gamma_{R_M}$ | 1 / Duration of isolation among mild symptomatic cases, remaining time to recovery (days<sup>-1</sup>) |
| $\alpha_{I_S}$ | 1 / Duration of infectiousness among severe cases, time to hospitalization (days<sup>-1</sup>) |
| $\gamma_H$ | 1 / Length of hospital stay (time until recovery or death) (days<sup>-1</sup>) |
| $\gamma_{\bar{H}}$ | 1 / Remaining time until recovery or death among hospital overflow patients<sup>-1</sup> (days<sup>-1</sup>) |
| $m_H$ | Case fatality ratio among hospitalized cases (HFR) |
| $m_{\bar{H}}$ | Case fatality ratio among hospital overflow patients |
| $k_n$ | Proportion of all contacts that happen with individuals from adjacent counties (as opposed to within the county) |
| $C$ | Hospitalization capacity, may be constant or vary over time representing capacity increase intervention |
| $E_0$ | Number of exposed individuals statewide at the time of epidemic onset |

Our transmission model does not include effects of vaccination, which began on December 14th, 2020. Initial vaccine deployment in Connecticut prioritized residents of congregate settings and individuals in the age group 75 years old and above. According to the CDC, COVID-19 vaccines achieve their full effectiveness 14 days after the second dose in a 2-dose series or a single dose in a 1-dose series [48]. As of March 1, 2021, about 5% of non-congregate population in Connecticut were fully vaccinated. Given that some of these people may have already experienced SARS-CoV-2 infection, that the majority of them were older people who do not mix as much as younger working-age population, and that the proportion vaccinated is within the uncertainty bounds of the cumulative incidence, vaccine effects before March 1, 2021 are unlikely to have a substantial impact on model projections. At the same time, some of the vaccine effects, such as incidence and case fatality reduction among older people are captured in the dynamics of time-varying model parameters. As vaccination coverage among young and working-age individuals increases, it will become important to incorporate vaccine effects in the transmission model.

## Time-varying model parameters

### Transmission rate *β*

Social distancing practices may reduce the value of the transmission rate *β*. We use data on close interpersonal contact in Connecticut for the entire duration of the modeling period [43], and assume the following functional form for the transmission rate:

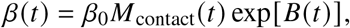

where *M*_contact_(*t*) is a smoothed normalized measure of close interpersonal contact at time *t* relative to the pre-epidemic level (February 1st - March 12th, 2020), and exp[*B*(*t*)] is a function that approximates residual changes in transmission parameter *β* that are not explained by changes in close contact and other time-varying parameters. Here, *B*(*t*) is a smooth function obtained by applying spline smoothing on a piecewise linear function *B*^∗^(*t*), where *B*^∗^(*t*) is modeled with 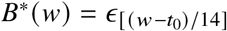 defined on bi-weekly knots *w* = {*t*_0_, *t*_0_ + 14, *t*_0_ + 28, …} over the observation period and linearly imputed between the knots. The Supplement shows plots of functions *M*_contact_(*t*) and *B*(*t*). We model the vector of random effects ***ϵ*** using a random walk of order one:

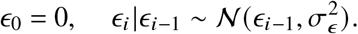

For the hyperparameter 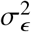, we use Inverse-Gamma(*a*_*ϵ*_, *b*_*ϵ*_) prior with a shape parameter *a*_*ϵ*_ = 2.5 and a rate parameter *b*_*ϵ*_ = 0.1. The function *B*(*t*) is also used to set *β*(*t*) in the future to test scenarios and potential intervention effects.

### Rates of isolation and recovery: 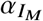 and *α*_*A*_

Widespread testing and contact tracing efforts can potentially reduce duration of infectiousness. While there is no information about the effectiveness of specific testing efforts implemented in Connecticut, the model accommodates the possibility of such reduction as a function of daily testing volume:

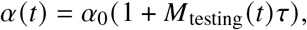

where *τ* is the size of testing effect per unit increase in testing volume measure *M*_testing_(*t*) modeled as:

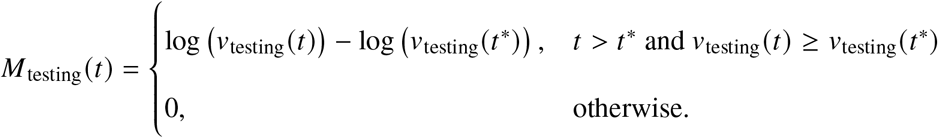

*v*_testing_(*t*) is a spline-smoothed measure of testing volume at time *t*. Testing efforts early in the epidemic were primarily used to confirm severe and highly symptomatic infections, and were unlikely to have any appreciable impact on overall duration of infectiousness. Early response daily testing volume is denoted by *v*_testing_(*t*^∗^). The Supplement shows the plot of function *M*_testing_(*t*). This approach is used to model time-varying rates 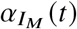 and *α*_*A*_(*t*) with 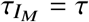 and *τ*_*A*_ = 0.5*τ*. The rate 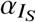 is assumed to remain constant over time.

### Severe fraction 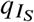

The probability of severe infection increases with age [49]. Age distribution of confirmed cases in the US has shifted toward younger people in the summer compared to spring [50]. We model the time-varying proportion of infections that are severe as:

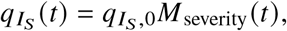

where measure of severity *M*_severity_(*t*) is a normalized spline-smoothed proportion of cases 60+ years old among all cases detected at time *t* relative to a baseline level. Since testing availability affects this proportion, we assume that *M*_severity_(*t*) = 1 for all *t < t*^∗^, where *t*^∗^ denotes the time when testing became widely available. The Supplement shows the plot of function *M*_severity_(*t*). Since 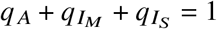, we also model *q* _*A*_ and 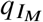 as functions of time.

### Rate of hospital discharge *γ*_*H*_

We estimate the time-varying rate of hospital discharge *γ*_*H*_ (*t*) (including deaths and alive discharges) as a reciprocal of the average length of hospital stay at time *t*, which has been inversely correlated with the incidence of COVID-19. The average length of hospital stay at time *t* is approximated using a spline-smoothed monthly averages of this quantity and is provided in the Supplement.

### Hospital case fatality ratio *m*_*H*_

Overall and hospital case fatality ratios may vary over time for various reasons. We model HFR as:

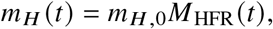

where *M*_HFR_(*t*) is a normalized spline-smoothed HFR at time *t* relative to the baseline HFR = *m*_*H*, 0_. HFR is calculated as a ratio of hospital deaths at time *t* to hospital admissions at time (*t* − HLOS(*t*)), where HLOS(*t*) is an average hospital length of stay at time *t*. The Supplement shows the plot of function *M*_HFR_(*t*).

## Model calibration and Bayesian posterior inference

We calibrate the posterior distribution of model parameters to estimated hospitalizations and hospital deaths coming from non-congregate settings using a Bayesian approach. Sampling from the joint posterior distribution of calibrated model parameters is performed using Markov Chain Monte Carlo. The Supplement provides detailed description of the data likelihood, calibration approach, sampling algorithm, implementation, and results, including convergence and estimated joint posterior distribution of model parameters, as well as description of prior distributions along with data sources. Source code and aggregated data used to produce model projections are available from https://github.com/fcrawford/covid19_ct. To generate model projections, we sample from the joint posterior distribution of estimated parameters, simulate transmission dynamics for a given combination of parameters, and compute pointwise averages (means or medians) and posterior predictive intervals for each time point.

### Ethics statement

This analysis does not use patient-level data with personal identifiers.

## Results

### Estimates of epidemiologic features

Figure 3 shows the results of model calibration along with estimates of important epidemiologic parameters for the period between March 1, 2020 - March 1, 2021. Plots of hospitalizations, effective reproduction number (*R*_eff_), and cumulative incidence are overlaid with the dates of Governor’s interventions, reopening of schools and universities in the fall and winter, and the starting date of vaccination campaign. Model fit to observed dynamics of hospitalizations and deaths shows that data points track with mean projections and largely fall within uncertainty intervals.

**Figure 3:**
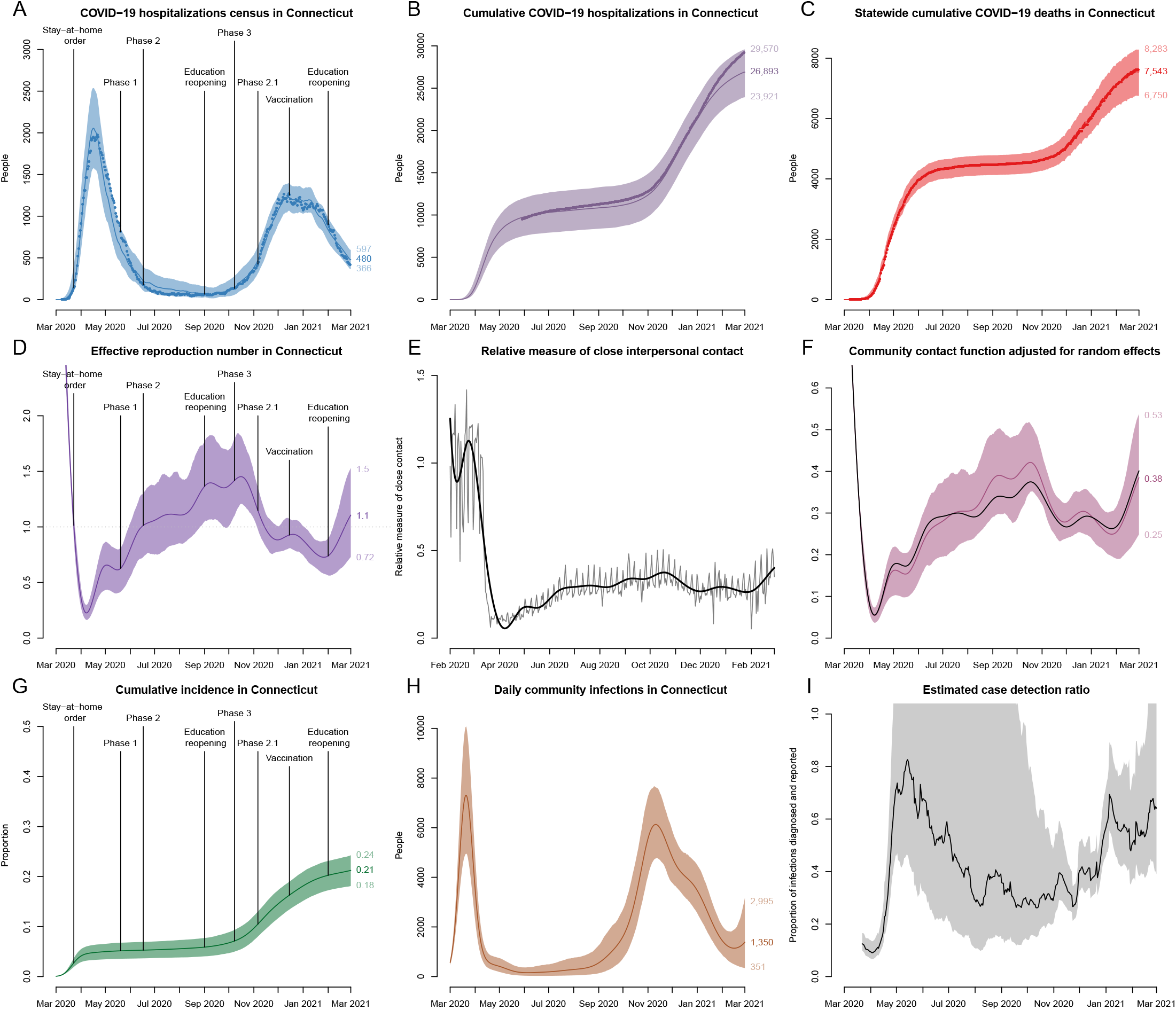
Model fit to observed data and estimates of epidemiologic features of SARS-CoV-2 transmission in Connecticut. Top row shows calibration results for: A: observed COVID-19 hospitalizations census, B: cumulative hospitalizations and C: cumulative deaths in Connecticut. Observed time series are shown as points and correspond to total hospitalizations and deaths among all Connecticut residents. The model is calibrated to estimated data series coming from non-congregate settings, and model projections are adjusted by the estimated difference to reflect the totals for congregate and non-congregate settings. Middle row: D: effective reproduction number, E: normalized measure of close contact relative to the pre-epidemic period along with the spline approximation (thick solid line), and F: contact function adjusted for estimated random effects that capture residual variation in transmission that is not explained by dynamics of close contact and other time-varying parameters. For comparison, black line in plot F shows smoothed normalized close contact metric unadjusted for random effects. Bottom row: G: cumulative incidence of SARS-CoV-2 infection, H: daily new infections, and I: estimated case detection ratio in non-congregate settings in Connecticut. Solid lines represent model-projected means and shaded regions represent 95% posterior predictive intervals. Some of the plots are overlaid with the key intervention dates (lockdown and phased reopening), as well as important event dates, including reopening of schools and universities and beginning of vaccination campaign.

We estimate that *R*_eff_ dropped substantially in mid-March and remained below one through mid-June. For the rest of the summer, mean estimated *R*_eff_ was slightly above one consistent with low numbers of case counts and hospitalizations in the summer. A major increase of *R*_eff_ started in mid-August and continued with the reopening of schools and colleges. It reached a maximum mean value of 1.45 by mid-October, followed by a slow decline through the rest of the year. A second increase in *R*_eff_ started at the end of January 2021 as schools and universities reopened for the spring semester. The dynamics of *R*_eff_ follows closely the dynamics of close contact. Plot F in Figure 3 shows that the estimated dynamics of transmission parameter captured by a community contact function (measure of close contact adjusted for estimated random effects) exhibits small deviations from the measure of close contact. Our estimate of *R*_0_ is 4.7 (95%CI: 4.3 – 5.1), consistent with estimates reported elsewhere [51, 52]. However, this estimate depends on assumptions about initial conditions and is not identifiable from the shape of early exponential increase alone.

We estimate that cumulative incidence at the beginning of June 2020 was 5.2% (95%CI: 3.6 – 6.8%) consistent with the results of community-based seroprevalence surveys conducted in Connecticut between April and June [53, 54] and other local modeling efforts that included Connecticut [55]. We estimate that as of March 1, 2021, cumulative incidence in Connecticut was about 21% (95%CI: 18 – 24%), which implies a cumulative case detection ratio of 36% (95%CI: 26 - 54%). Our estimates suggest that the case detection ratio varied substantially over time in a way that is not explained by the volume of PCR testing alone (plot I in Figure 3). An increase up to 80% in mid-May may be due to delayed testing, including postmortem diagnosis of first epidemic wave cases. The second spike in estimated case detection ratio in early January may be a consequence of testing and reporting disruptions related to the Christmas and New Year holidays.

We estimate the cumulative infection fatality ratio (IFR) to be 1.06% (95% CI: 0.93 – 1.24%) and the cumulative infection hospitalization ratio (IHR) to be 4.1% (95% CI: 3.6 – 4.7%). These estimates are based on data from all recorded deceased and hospitalized individuals, including those residing in congregate settings. Estimated IFR and IHR among residents of non-congregate settings are 0.43% (95% CI: 0.38 - 0.50%) and 3.4% (95% CI: 3.0 – 4.0%) respectively, which are somewhat lower than previously estimated in Connecticut [56], but are consistent with studies conducted elsewhere [6, 14, 49, 57].

### Predictive performance of the model

We illustrate predictive performance of the model by calibrating it to data up to several time points and producing projections of hospitalizations and deaths for the next two months. Projections into the future are simulated by propagating the latest available values of time-varying parameters into the future. A forecasting time horizon of two months is shown. Longer-term projections were less useful due to anticipated changes in policy, public behavior, and time-varying parameters. The time points were selected based on important events, such as reopening phases, as well as distinct stages of the epidemic. Figure 4 shows calibrations results, projections and actual data over the prediction period for the following calibration cut-off dates: May 20, 2020 (Phase 1 of reopening), June 17, 2020 (Phase 2 of reopening), September 1, 2020 (reopening of schools and colleges), October 15, 2020 (early indications of resurgence), and December 15, 2020 (early indications of the curve flattening during the second epidemic wave).

**Figure 4:**
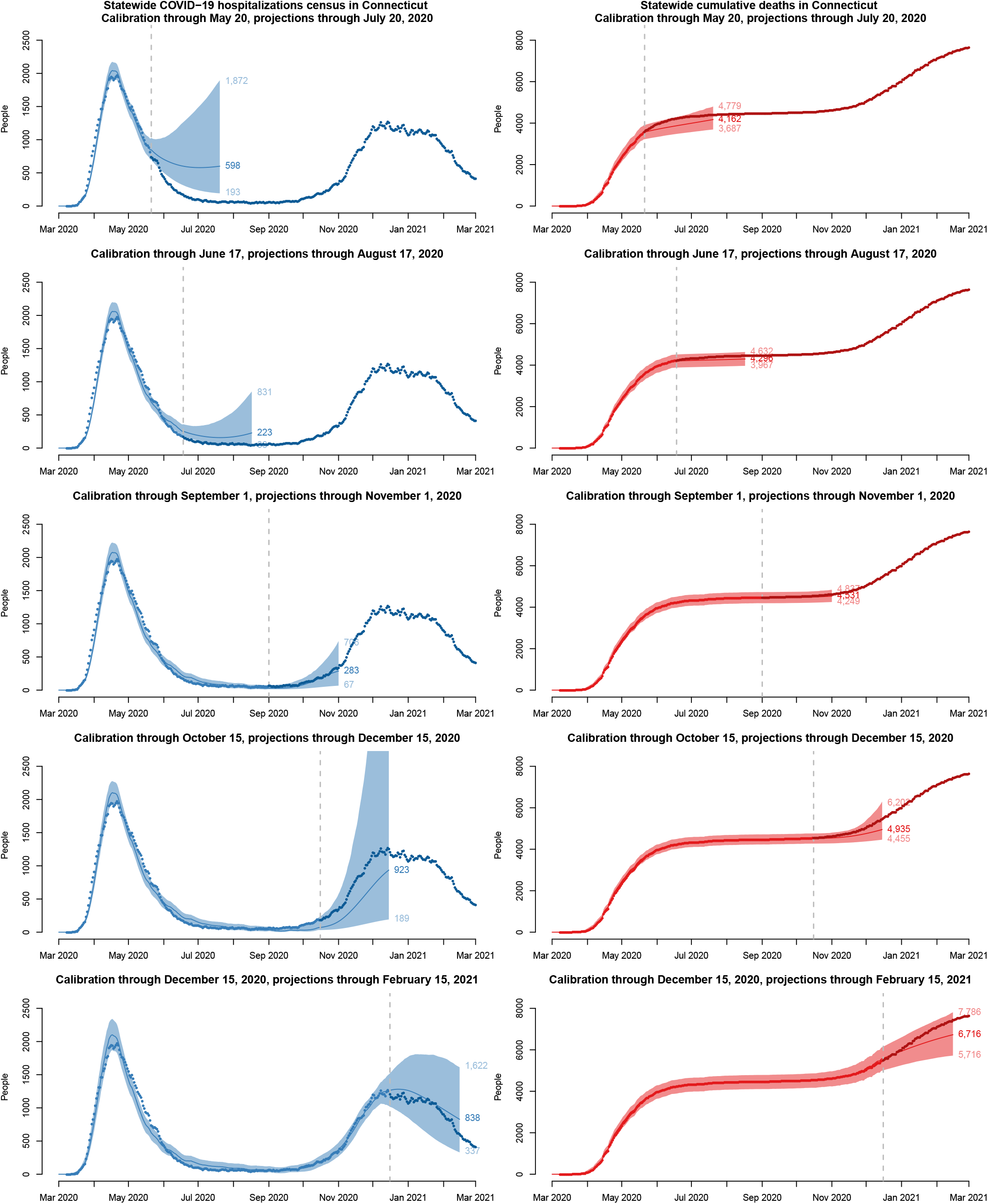
Posterior predictive performance of the transmission model calibrated using data up to the dashed line shown in each plot and projected forward for a period of two months. Solid lines represent model-projected means and shaded regions represent 90% posterior predictive intervals. Observed data are shown as points; lighter color points correspond to the data used in calibration.

These results show that projections become more accurate as data accrue, and that uncertainty intervals are generally tighter during periods of epidemic decline, when the upper bound of uncertainty interval for projected *R*_eff_ is below one. In early stages, our projections predicted higher hospitalization census compared to what was observed during summer 2020. The main reason for this mismatch is a sharp decline in severe infections proportion that was observed after the initial epidemic wave subsided, as well as the decline in the average length of hospital stay among hospitalized COVID-19 patients (plots H and F in Figure 1).

## Discussion: transmission modeling in the COVID-19 pandemic

As COVID-19 pandemic emerged, policymakers in many parts of the US and across the globe had to make quick decisions under high uncertainty. Access to reliable information about both the current state of the local epidemic and likely future outcomes is an important foundation to support policymakers in this process. Elected officials and public health agencies already have access to near real-time information about COVID-19 testing, case counts, hospitalizations, and deaths. However, these data may not provide timely insight into the current and future dynamics of COVID-19 transmission. Mathematical modeling of infectious diseases offers one way to address these questions, but is subject to limitations that may be difficult to communicate to policymakers. During our year-long collaboration with CT DPH, we learned several lessons in the process of developing, deploying, calibrating, revising, and communicating the outputs of COVID-19 transmission model to policymakers in Connecticut.

### Transmission dynamics in congregate settings may bias model-based estimates

Observable features of the initial epidemic wave in Connecticut - hospitalizations and deaths - were dominated by residents of skilled nursing and assisted living facilities [29]. In our initial efforts to model outcomes under the state’s stated reopening plans in May of 2020, we did not isolate transmission in congregate settings and treated all hospitalizations and deaths as those arising from a homogeneous mixing process within Connecticut population [27]. Transmission dynamics in congregate settings among higher risk individuals violate the homogeneous mixing assumption underlying compartmental modeling approaches. Merging cases, hospitalizations and deaths from congregate and non-congregate settings may bias estimates of *R*_eff_ and result in over-estimation of cumulative incidence, transmission potential, and infection fatality ratio. Recognizing this, we modified the model to only include residents of non-congregate settings.

### Detailed local data are necessary to capture time-varying epidemic features

Similar to many early transmission models that only used data from the first epidemic wave [13–16, 19], the first version of our model did not incorporate any temporal variation in the input parameters and included constant effects of school closure and lockdown [27]. However, as the data accrued, constant parameter values failed to achieve a good fit to the observed data. Our collaboration with CT DPH allowed us access to detailed local data informing time-varying model parameters. Incorporating time trends in important epidemic features like close interpersonal contact, risk profile of incident cases, hospital length of stay, and hospital case fatality ratio substantially improved model fit to observed data and its predictive performance. One of the limitations of our model is that it does not incorporate vaccination effects. However, during the modeling period ending on March 1, 2021 these effects are likely negligible in non-congregate residents due to low overall coverage that is primarily driven by an older age group, and are partially captured in the dynamics of time-varying parameters. As vaccination coverage increases, it will be important to include vaccination effects in the transmission model.

### Anticipated changes in time-varying epidemic features warrant scenario analysis

The most substantial changes in time-varying model parameters occurred as the first epidemic wave began to subside in early summer 2020. Figure 4 illustrates modest predictive performance of our model during early stages. In this example, future projections are made by propagating the last observed value of time-varying model parameters into the future. When substantial changes in epidemic features are anticipated in the future, scenario analysis may offer a better way to represent the true uncertainty.

### Close interpersonal contact drives transmission

One of the most important data sources that substantially improved our model performance is a measure of close interpersonal contact based on mobile device data. This is a novel metric, whose dynamics exhibit different behavior from that observed in several publicly available mobility metrics, such as maximum distance traveled or time spent away from home, which mostly returned to their baseline values by mid-summer 2020 [43]. Combined with other time-varying parameters, the close contact metric captured most of the variation in transmission over time. While our approach to use random effects to capture residual variation in transmission dynamics proved feasible and useful, the shape of the resulting contact function exhibits small deviations from the dynamics of close interpersonal contact (plot F in Figure 3).

### Timing of interventions may not always reflect timing of behavioral changes

Public perception of risk appears to be an important time-dependent confounder that is hard to measure or predict. Behavior changes that drive transmission, such as social distancing or mask wearing, may not coincide in time with respective interventions. In Connecticut, close interpersonal contact dropped substantially before the lockdown order went in effect and started rebounding before it was lifted. Many transmission models [6, 15, 16], including the early version of our model [27] assume constant intervention effects that modify a given set of parameters at the time of their enactment. Measuring critical features of transmission directly improves model-based projections and offers insights into the estimation of intervention effects.

### Case counts may be an unreliable proxy for infections

Policymakers often rely on case counts and test positive proportion as direct measures of infection incidence. Even though dynamics of detected cases may be a reasonable qualitative indicator of disease incidence, we found that its usefulness for modeling purposes is limited. It is widely recognized that detected case counts depend on the underlying infection incidence and testing volume [24, 58], however we found that changes in testing strategies and human behavior may lead to non-monotonic relationship between testing volume and case detection rate. Since case counts dynamics are usually widely available across geographic locations, and are often among a few data sources easily accessible by researchers, many nationwide models rely on case counts, possibly adjusting for testing volume [7, 22]. Our experience shows that this approach may be misleading. We therefore chose to rely on hospitalizations and death data for model calibration as these epidemic features are less susceptible to unmeasured time-varying confounding.

### Incorporating diverse data sources improves credibility of model-based inferences

Our modeling projections have been one of the many analytic products informing policy response in Connecticut along with surveillance data on case counts, testing volume and test positive proportion, trends in emergency department visits presenting symptoms of COVID-like illness, outbreak investigations, wastewater monitoring [28], and trends in close interpersonal contact [43] among others. For a variety of reasons, it can be difficult for public health decision-makers to enact policy responses based on predictions about the future from transmission models. However, when model projections tell a cohesive story in combination with other analytic products, they may fill in missing pieces and offer insights into the current and future epidemic dynamics.

## Conclusions and limitations

Our year-long effort to provide policymakers with predictions of COVID-19 dynamics in Connecticut showed that standard SEIR-type transmission models work best in large well-mixing populations with constant transmission rates and without major external shocks, like interventions or importation of infections. In order to make valid inferences, it is necessary to incorporate temporal changes in model parameters reflecting interventions, human behavior, heterogeneity in mixing patterns, and measurement errors. Given geographic heterogeneity in the dynamics of these features, models that incorporate contextual information and local data are needed to support local policymakers. While modeling approach described in this paper captures many important features of epidemic dynamics that are often omitted in simpler SIR-type models, our model could be extended to reflect more granular geographic variation, age structure and contact patterns between different age groups, vaccination effects, and other time-varying features of COVID-19 epidemic. When it works best, modeling provides heuristics, guidance, and what-if scenarios for the future offering insights that are otherwise unavailable.

## Data Availability

All data and code used in this analysis are available from https://github.com/fcrawford/covid19_ct.

## Acknowledgements

We are grateful to Jacqueline Barbieri, Maciej F. Boni, Jessica Brockmeyer, Jared Campbell, Matthew Cartter, Alexandra Edmundson, Hanna Ehrlich, Deidre Gifford, Sydney A. Jones, Edward H. Kaplan, Patrick Kenney, Alison Kleppinger, Albert Ko, Terry Rabatsky-Ehr, Olivia Schultes, Lynn Sosa, and Thomas Valleau. We thank Whitespace Solutions Ltd for providing the contact data. We thank the Connecticut Hospital Association for providing data on COVID-19 hospitalizations and deaths. We thank Connecticut Department of Public Health for providing detailed data on testing, cases and deaths. We thank Stony Brook Research Computing and Cyberinfrastructure, and the Institute for Advanced Computational Science at Stony Brook University for access to the high-performance SeaWulf computing system.

## Author contributions

Conceptualization: FWC, OM; Methodology Development: FWC, OM, ZRL; Software Development: FWC, OM, ZRL; Validation: OM; Formal Analysis: OM, ZRL; Resources: FWC; Data Curation: FWC, OM; Writing – Original Draft: OM; Writing – Review & Editing: FWC, OM, ZRL; Visualization Preparation: FWC, OM, ZRL; Supervision Oversight: FWC; Project Administration: FWC, OM; Funding Acquisition: FWC.

## Supplementary Material: Detailed Methods

### Estimating hospitalizations and hospital deaths coming from non-congregate settings

Our transmission model aims to capture disease spread in the community and excludes residents of congregate settings, whose contact patterns violate homogeneous mixing assumption. Available hospitalizations data do not disaggregate by the patient’s place of residence at the time of diagnosis or hospitalization. We have therefore estimated the time-varying number of hospital admissions and hospitalizations census that came from congregate settings and excluded them from the observed time series. We received data on daily COVID-19 death counts in hospitals disaggregated by the type of residence (congregate vs. non-congregate) at the time of diagnosis or hospitalization from Connecticut Department of Public Health (CT DPH). Posterior distribution of model parameters was calibrated to the estimated time series of cumulative hospitalizations and hospitalizations census coming from non-congregate settings and to the observed time series of hospital deaths in this population.

The time-varying proportion of hospitalization census and cumulative hospitalizations coming from congregate settings was estimated as follows:

1. We first estimated the cumulative number of hospitalizations coming from congregate settings as the total number of hospital deaths among residents of congregate settings divided by the hospital case fatality ratio (HFR) among this population, and adjusting for an average hospital length of stay. The estimated HFR among residents of congregate settings in Connecticut is 0.38 and was obtained from the CT DPH based on a survey of a sample of 50 nursing homes, representing about a quarter of all nursing homes in Connecticut [29].
2. Next, we computed the cumulative proportion of hospitalizations coming from congregate settings as an estimate of the number of cumulative hospitalizations coming from congregate settings divided by the total cumulative number of hospitalizations as of the same date.
3. The proportion of hospital admissions coming from congregate settings varied over time. To approximate temporal dynamics of this proportion, we assumed that it follows the same pattern as the time-varying proportion of deaths among hospitalized residents of congregate settings among all hospital deaths:

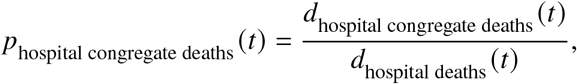

where *d*(*t*) denotes smoothed daily death counts calculated as a first order difference of spline-smoothed cumulative counts of respective time series. We then re-scaled this time-varying proportion relative to the cumulative proportion of deaths among hospitalized residents of congregate settings among all hospital deaths and lagged it back by the average hospital length of stay. The resulting time-varying function was then multiplied by the cumulative proportion of hospitalizations coming from congregate settings estimated in step 2 to obtain a time-varying estimate of the proportion of hospital admissions coming from congregate settings at time *t*.

This time-varying proportion was then applied to the CHA-reported daily hospital census and daily hospital admissions to estimate the number of current hospitalizations, daily hospital admissions, and cumulative hospitalizations coming from congregate settings, and respective time series coming from non-congregate settings follow directly.

### Data smoothing for time-varying parameters

All time-varying parameters described in detail in the Methods section rely on smoothed functions of observed data. Figure 5 shows five functions of observed data series overlaid by a smooth function (red line), which is used as a component of time-varying model parameters.

**Figure 5:**
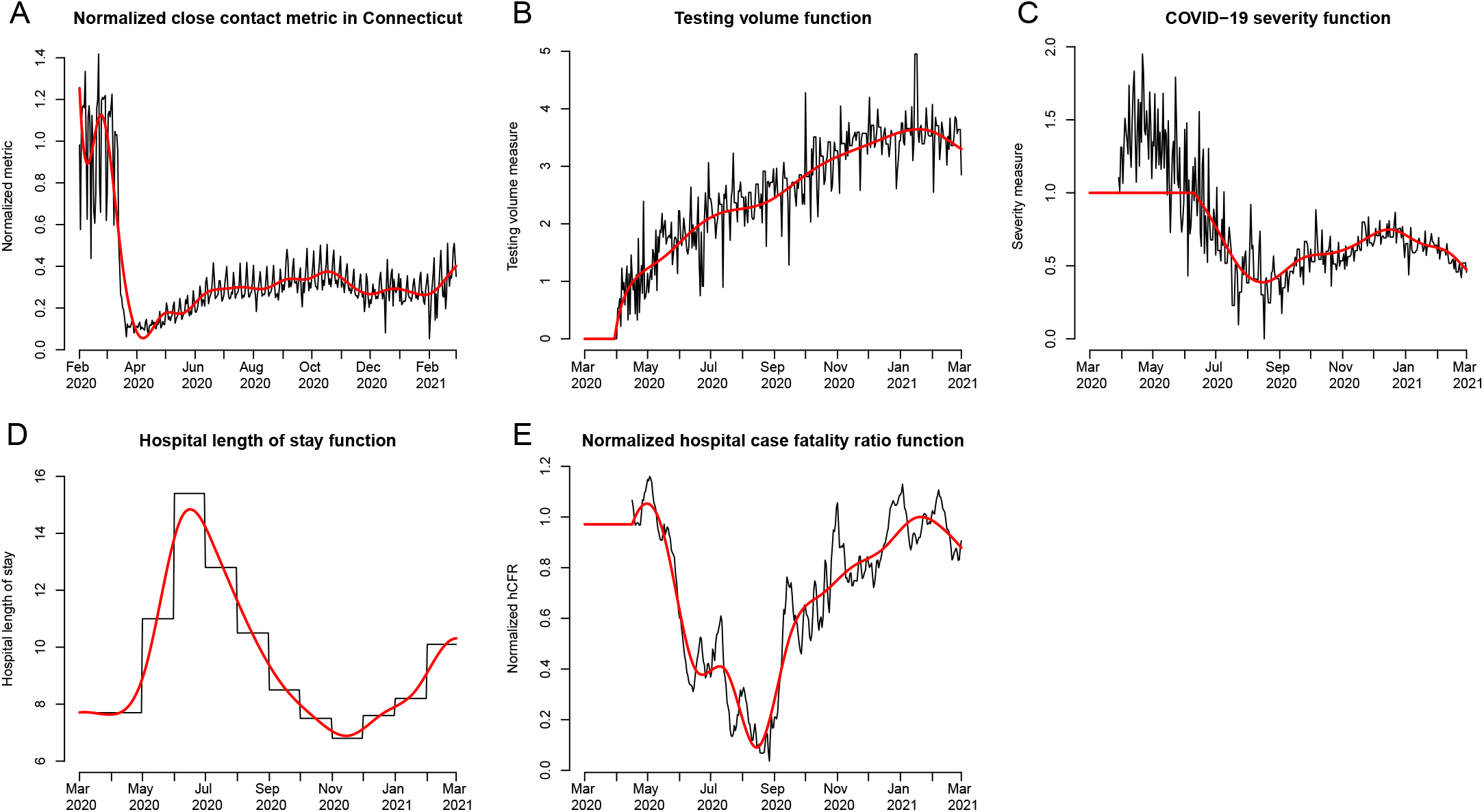
Smooth functions of observed data used as components of time-varying model parameters. A: normalized close contact metric, function *M*_contact_ (*t*); B: testing volume function *M*_testing_ (*t*); C: COVID-19 severity function, normalized proportion of cases 60+ years old among all cases detected at time *t*, function *M*_severity_ (*t*); D: average hospital length of stay among COVID-19 patients, reciprocal of rate of hospital discharge *y*_*H*_ (*t*); E: normalized hospital case fatality ratio, function *M*_HFR_ *t*. Black lines show input data trajectories and red lines show smooth functions of observed data.

### Model calibration and Bayesian posterior inference

We calibrate the posterior distribution of model parameters to the estimated hospitalizations and hospital deaths coming from non-congregate settings using a Bayesian approach with a Gaussian likelihood. Model-based estimates of observed quantities are adjusted for reporting lags. The distributions of hospitalizations census, cumulative hospitalizations, and hospital deaths are given by:

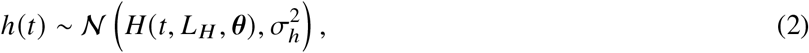

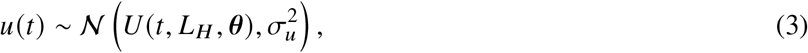

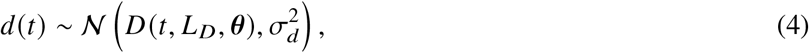

where *H*(*t, L*_*H*_, *θ*), *U*(*t, L*_*H*_, *θ*), and *D* (*t, L*_*D*_, *θ*) are model-projected hospitalizations census (lagged by *L*_*H*_), cumulative hospitalizations (lagged by *L*_*H*_), and cumulative deaths (lagged by *L*_*D*_) at time *t* under parameter values *θ*. Prior distributions imposed on calibrated model parameters ensure non-negative values of model compartments.

We put uniform priors on *L*_*H*_ and *L*_*D*_ over a range of plausible integer values. Reporting lags are correlated with other unknown parameters, including latency period, time between infection and hospitalization, time between infection and death, and length of hospital stay, therefore *L*_*H*_ and *L*_*D*_ should not be strictly interpreted as reporting lags. We put the same independent Inverse-Gamma(*a, b*) prior on all three hyperparameters 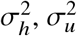, and 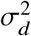 with *a* = 0.5 and *b* = 5 × 10^6^ at the end of the modeling period. The prior was gradually relaxed as observed time series increased. For the purposes of future forecasting, as opposed to estimation of epidemiologic features from the past model dynamics, we imposed a tighter prior on these hyperparameters to achieve reasonable posterior predictive intervals beyond the observation period.

We construct the posterior distribution over unknown parameters (*θ, σ*) as:

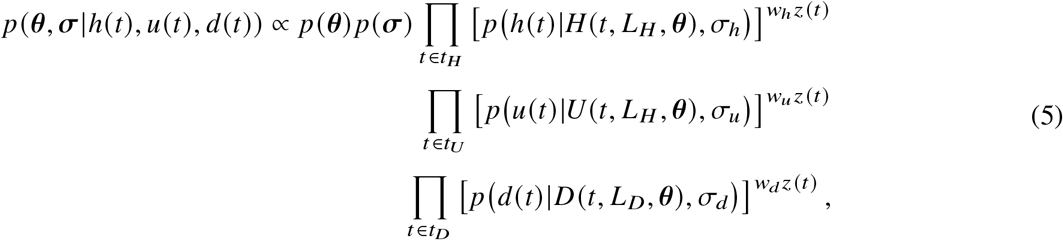

where 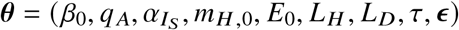 and *σ* = (*σ*_*h*_, *σ*_*u*_, *σ*_*d*_, *σ*_*ϵ*_).

We assume the date of epidemic onset to be February 16th, 2020 - 21 days before the first case was officially confirmed in Connecticut on March 8th, 2020, and initialize the model with *E*_0_ exposed individuals at the time of epidemic onset, setting the size of all downstream compartments to be zero. County-level distribution of *E*_0_ is fixed and was estimated based on the county population size and dates of first registered case and death in each county.

Each likelihood term is weighted by the time-dependent weight *z*(*t*) times the weight assigned to a respective time series. We let the weight function *z*(*t*) take the following form,

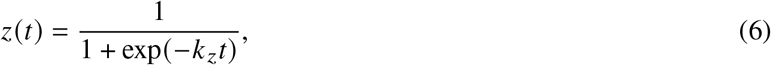

where *z*(*t*) is the weight assigned to an observation at time *t, t* ∈ {*t*_0_, …, *t*_max_}, and the correspondence between {*t*_0_, …, *t*_max_} and calendar time is set such that *t* = 0 corresponds to 90 days prior to the most recent observation. Parameter *k*_*z*_ controls the smoothness of logistic function. We set *k*_*z*_ = 0.01, resulting in a range of weights between 0.06 − 0.7 for the duration of observation period.

We set *w*_*h*_ = 0.89, *w*_*u*_ = 0.01, and *w*_*d*_ = 0.1. We place a large weight on the hospitalizations census, since this time series is most sensitive to changes in epidemic dynamics, and a small weight on cumulative hospitalizations, since it measures a feature that is related to hospitalizations census. The range of observation times differ for different time series. For hospitalizations census and deaths, observation times start with the first non-zero observation. For cumulative hospitalizations, observation times start on May 29, 2020 when these data started being reported routinely.

### Posterior sampling

Sampling from the joint posterior distribution of (*θ, σ*) given in (5) is performed using Markov Chain Monte Carlo (MCMC). We employ a hybrid algorithm that combines elliptical slice sampling (ESS) [59], Gibbs sampling, and Metropolis-Hastings sampling with random walk proposals. We first provide an overview of the steps in drawing samples from the full posterior distribution and then describe each steps in more details. Let = (*θ*_MH_, *θ*_ESS_), where 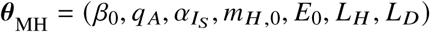 and *θ*_ESS_ = (*τ, ϵ*). The sampler proceeds with the following steps:

1. Update *θ*_ESS_ |*θ*_MH_, *σ* using ESS.
2. Update *θ*_MH_ |*θ*_ESS_, *σ* with a Metropolis-Hastings step.
3. Update hyperparameters *σ*|*θ* with a Gibbs sampler step.

**Update** *θ*_**ESS**_ |*θ*_**MH**_, *σ*: We use a rejection-free sampler (ESS) to sample a vector of random effects *ϵ* and a testing effect *τ*. The ESS operates by drawing samples from the ellipse defined by a Gaussian prior, and then accept or reject the samples by evaluating the likelihood component. Within the slice sampling step, the sampler moves along the generated ellipse and always accepts a new set of parameters.

**Update** *θ*_**MH**_ |*θ*_**ESS**_, *σ*: We implement a Metropolis-Hastings algorithm with random walk proposals for *θ*_MH_. Proposals for *L*_*H*_ and *L*_*D*_ are made on a subset of integers bounded by a prior distribution on lags. All other elements of *θ*_MH_ are continuous.

**Update hyperparameters** *σ*|*θ*: The hyperparameters 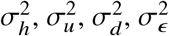 are updated with Gibbs sampler steps:

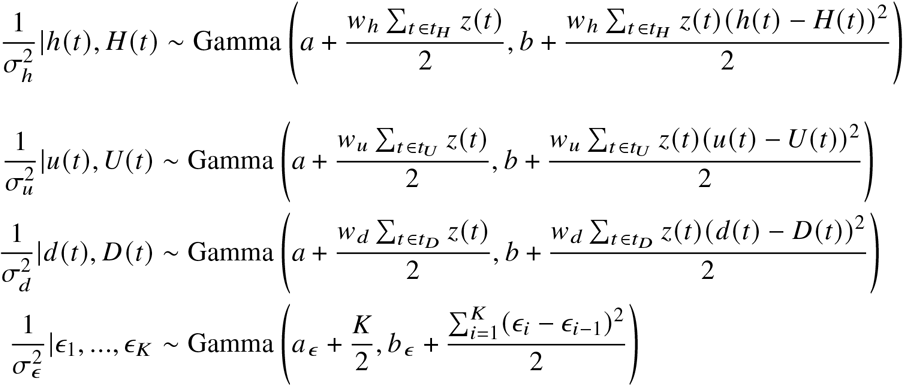

We ran 10 chains of the sampler for 22,000 iterations each, discarded the first 2,000 draws from each chain, and thinned each chain by a factor of 20. The final posterior sample combines the resulting thinned chains. Based on the visual inspection of individual parameter trace plots, we found that 10,000 iterations is sufficient for the chain to converge in practice. Figure 6 illustrates model calibration results and convergence. The top row shows each of 10 thinned MCMC sampler chains overlaid. The middle row shows posterior histograms, marginal means and 95% posterior credible intervals of calibrated model parameters. The bottom row shows calibrated random effects function *B*(*t*). To generate uncertainty intervals of model projections, we sample from the joint posterior over estimated parameters, and find pointwise 90% or 95% posterior predictive intervals for each time point.

**Figure 6:**
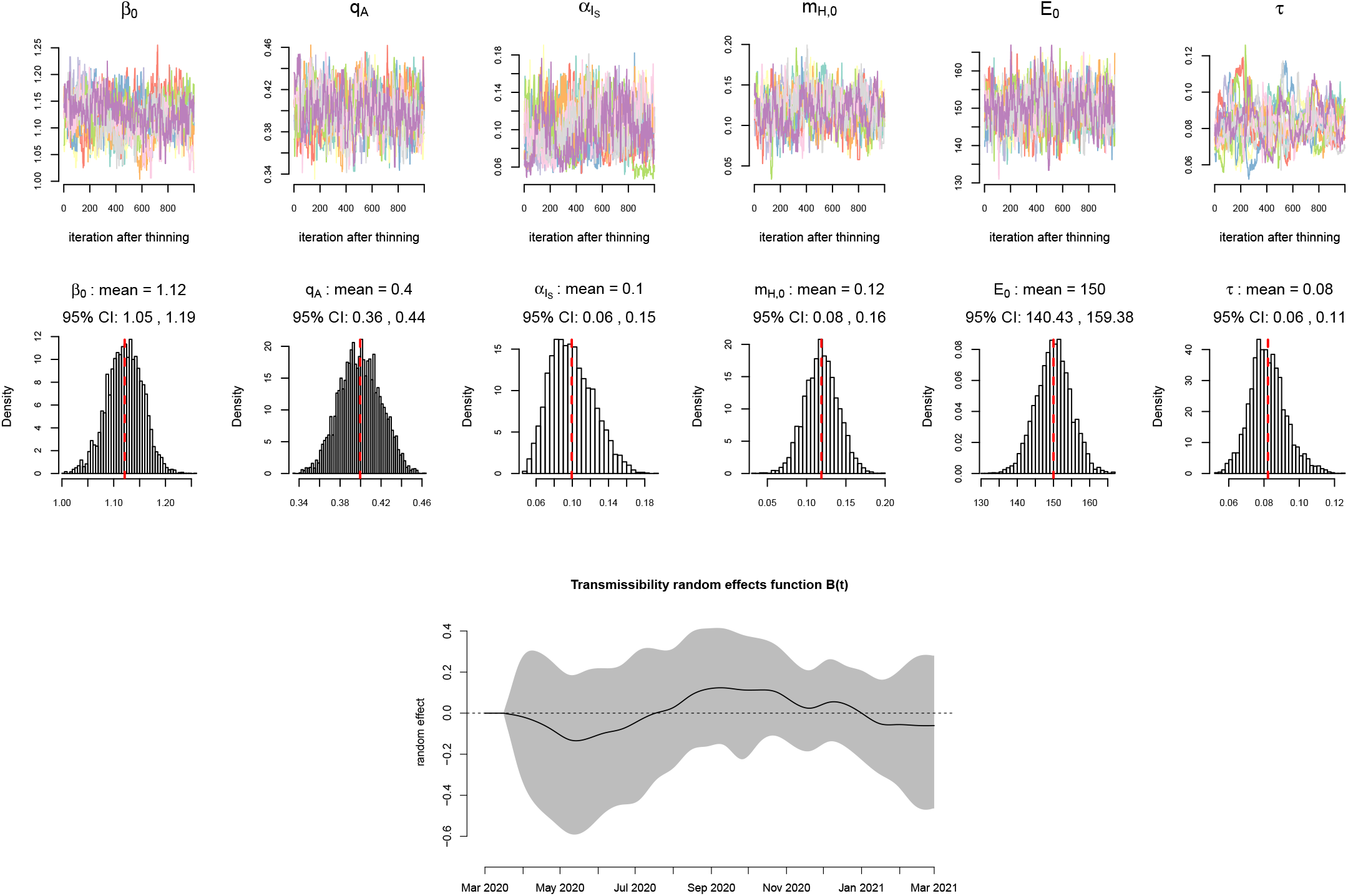
Model calibration results: trace plots of MCMC sampler chains, marginal posterior histograms of calibrated parameters, and random effects function *B t*. MCMC trace plots show 10 thinned chains plotted using different colors. Plot titles of posterior histograms include marginal posterior means and 95% credible intervals (CI). Dashed red lines correspond to posterior means of model parameters. In the transmissibility random effects function plot, black line shows the mean value of *B*(*t*) at each time point, and the shaded region shows 95% CI.

### Prior and posterior distributions of model parameters

Let *θ*_MH,CONT_ denote the subset of continuous transmission model parameters whose joint distribution is calibrated to observed data and updated with Metropolis-Hastings step: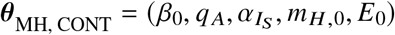. For each individual parameter *θ* ∈*θ*_MH,CONT_, we specify a fixed support [*θ*_min_, *θ*_max_], and put independent beta priors on the transformed parameter, i.e.,

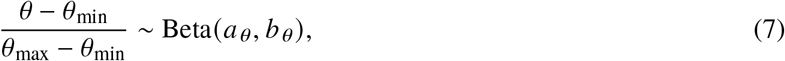

where the shape parameters *a*_*θ*_ and *b*_*θ*_ are set to let *θ* have mean *µ*_*θ*_ and standard deviation *σ*_*θ*._ Table 2 provides the summary of (*µ*_*θ*_, *σ*_*θ*_, *θ*_min_, *θ*_max_) for all parameters in *θ*_MH,CONT_ along with data sources. For parameters whose values were fixed, only the mean is given. Parameter *τ* is updated in the ESS step, therefore in calibration, we assume a Gaussian prior distribution on log(*τ*), whose mean and SD are shown in Table 2.

**Table 2:** Prior distributions of model parameters.

| Parameter | Mean | SD | Lower | Upper | Source |
| --- | --- | --- | --- | --- | --- |
| $q_A$ | 0.4 | 0.02 | 0.3 | 0.5 | [60–65] |
| $q_{IS,0}/(1 - q_A)$ | 0.07 | - | - | - | 7% of symptomatic infections are severe [4, 6, 49] |
| $\beta_0$ | 1 | 0.5 | 0.01 | 2.00 | A diffuse prior assumed |
| $\delta$ | 1/4 | - | - | - | [6, 12, 15, 23, 62, 66–73] |
| $\alpha_{A,0}$ | 1/7 | - | - | - | [66, 74–79] |
| $k_A$ | 0.5 | - | - | - | [23, 61, 76, 80, 81] |
| $\alpha_{IM,0}$ | 1/4 | - | - | - | [4, 6, 15]; of 4 days, 2 are assumed to be presymptomatic [23, 66, 67] |
| $\gamma_{RM}$ | 1/7 | - | - | - | [74, 78, 79, 82] |
| $\alpha_{IS}$ | 1/9 | 0.03 | 0.02 | 0.2 | [23, 66, 67, 83, 84] |
| $m_{H,0}$ | 0.13 | 0.05 | 0.01 | 0.25 | Estimated from Connecticut COVID-19 hospitalization data [41] |
| $m_{\bar{H}}/m_H$ | 1.5 | 0.25 | 1 | 2 | Assumed |
| $\log(\tau)$ | -2.5 | 0.12 | - | - | Assumed, Gaussian prior distribution |
| $k_n$ | 0.015 | - | - | - | Assumed |
| $E_0$ | 150 | 5 | 100 | 200 | Assumed |
| $L_H$ | - | - | 5 | 14 | Assumed, uniform prior between lower and upper values |
| $L_D$ | - | - | 5 | 14 | Assumed, uniform prior between lower and upper values |

Asymptomatic infections play an important role in transmission of SARS-CoV-2 [85–87], but estimates of the proportion of infections that do not exhibit symptoms vary substantially [60, 61, 88]. The true proportion of asymptomatic infections is important for projections and policy planning due to its relationship to evolving herd immunity. Reported estimates of asymptomatic proportion range between 6 – 96%, and the authors of a recent review recommend a range between 40 – 45% [60]. Another review reported an overall asymptomatic proportion estimate of 20%, and 31% among the studies that included follow-up [61]. Large population-based studies conducted in Spain and in the U.K. provide estimates of asymptomatic proportion between 22-36% [89, 90]. We calculated age-adjusted weighted average of several estimates available from the literature: Nishiura et al. [63] estimated 30.8% among Japanese citizens evacuated from Wuhan, China. We applied this estimate to age group 20-64 years old. Mizumoto et al. [64] estimated 17.9% among infections on the Diamond Princess cruise ship. We applied this estimate to age group 65 plus years old, which is the age group, in which most infections occurred. We assumed 65% for age group 0-19 years old, consistent with findings reported by Russell et al. [65], where 4 out of 6 infections in this age group were asymptomatic among passengers of the Diamond Princess. The average was weighted by the age distribution of Connecticut population, resulting in the estimate of *q* _*A*_ = 0.37, which is consistent with systematic reviews and population-based studies. At the time of this writing, the best estimate of asymptomatic proportion recommended by the CDC was 40% [62]. We center the prior distribution of *q*_*A*_ at 0.4 and allow it to vary between 0.3 and 0.5. The proportion of severe cases early in the epidemic 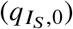 is assumed to be 7% of symptomatic infections in line with [4, 6, 49]. Combined with the estimates of asymptomatic proportion, this translates into an estimate of severe proportion of 4.2% (3.5-4.9%) of all infections.

Duration of latency (1/*δ*) was fixed, since consistent estimates of this parameter are available from the literature, and because it is highly correlated with several calibrated parameters, including reporting lags and parameters that determine duration of infectiousness and time between infection and recovery or death. We assume an average of 4 days of latency [6, 15, 23] and 2 days of presymptomatic infectiousness [23, 66, 67] resulting in an average incubation period of 6 days consistent with [12, 62, 68–73].

Parameters 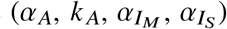 collectively determine the force of infection at any given time. Force of infection, transmission parameter *β* and initial number of exposed individuals *E*_0_ together determine the early growth of the epidemic. Without additional data, all of these parameters cannot be simultaneously identified. We therefore fixed the values for a subset of these parameters based on available estimates and assumed a diffuse prior on *β*, which absorbs additional variation of parameters that determine the force of infection.

Duration of infectiousness of asymptomatic individuals is unknown, but is likely shorter than that of symptomatic individuals [74–77]. While several studies estimated that viral RNA could be detected in upper respiratory tract for 2-3 weeks [91, 92], findings from [74, 78, 79] show that in mild-to-moderate symptomatic patients live virus could be isolated for a substantially shorter time period: up to 7-10 days from the day of symptom onset, suggesting the duration of infectiousness of up to 12 days. Based on these estimates and [66], we assume the duration of infectiousness of 7 days among asymptomatic individuals. Although multiple studies have shown similar viral loads among symptomatic, presymptomatic and asymptomatic cases [75, 93, 94], evidence suggests that asymptomatic individuals are less infectious that symptomatic, likely due to higher viral shedding while coughing and longer duration of infectiousness among symptomatic individuals [61, 74, 76, 95, 96]. Multiple estimates of relative infectiousness of asymptomatic individuals compared to symptomatic are available and range from close to zero to above one [23, 61, 76, 80, 81]. Generally, estimates that are based on attack rates are lower than those based on viral shedding or time until the first negative PCR test. In our analysis, we set relative infectiousness of asymptomatic cases to be 0.5 [23, 61, 80].

Although the duration of infectiousness of mild-to-moderately symptomatic cases may be up to 12 days [74, 78, 79], we assume that the majority of symptomatic individuals self-isolate shortly after developing symptoms. We set the duration of infectiousness of symptomatic cases to be 4 days [4, 6, 15], 2 of which are assumed to represent presymptomatic infectiousness [23, 66, 67]. We further assume that the duration of self-isolation until recovery is 7 days [74, 78, 79].

We calibrate the rate of hospitalization among severe cases 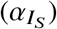, assuming a mean of 9 days between the onset of infectiousness and hospitalization: 2 days of presymptomatic infectiousness plus 7 days between the onset of symptoms and hospitalization [83, 84].

## References

[1] Neil M Ferguson, Daniel Laydon, Gemma Nedjati-Gilani, Natsuko Imai, Kylie Ainslie, Marc Baguelin, Sangeeta Bhatia, Adhiratha Boonyasiri, Zulma Cucunubá, Gina Cuomo-Dannenburg, Amy Dighe, Ilaria Dorigatti, Han Fu, Katy Gaythorpe, Will Green, Arran Hamlet, Wes Hinsley, Lucy C Okell, Sabine van Elsland, Hayley Thompson, Robert Verity, Erik Volz, Haowei Wang, Yuanrong Wang, Patrick GT Walker, Caroline Walters, Peter Winskill, Charles Whittaker, Christl A Donnelly, Steven Riley, Azra C Ghani, and Imperial College COVID-19 Response Team. Report 9: Impact of non-pharmaceutical interventions (NPIs) to reduce COVID-19 mortality and healthcare demand. 2020. doi: https://doi.org/10.25561/77482. URL https://www.imperial.ac.uk/media/imperial-college/medicine/sph/ide/gida-fellowships/Imperial-College-COVID19-NPI-modelling-16-03-2020.pdf.

[2] David Adam. Special report: The simulations driving the world’s response to COVID-19. Nature, 580(7803): 316, 2020.

[3] Centers for Disease Control and Prevention. COVID-19 Mathematical Modeling. https://www.cdc.gov/coronavirus/2019-ncov/covid-data/mathematical-modeling.html, Updated Aug. 7, 2020.

[4] Stephen M Kissler, Christine Tedijanto, Edward Goldstein, Yonatan H Grad, and Marc Lipsitch. Projecting the transmission dynamics of SARS-CoV-2 through the postpandemic period. Science, 368(6493):860–868, 2020.

[5] Seth Flaxman, Swapnil Mishra, Axel Gandy, H Juliette T Unwin, Thomas A Mellan, Helen Coupland, Charles Whittaker, Harrison Zhu, Tresnia Berah, Jeffrey W Eaton, Mélodie Monod, Imperial College COVID-19 Response Team, Azra C. Ghani, Christl A. Donnelly, Steven Riley, Michaela A. C. Vollmer, Neil M. Ferguson, Lucy C. Okell, and Samir Bhatt. Estimating the effects of non-pharmaceutical interventions on COVID-19 in Europe. Nature, 584(7820):257–261, 2020. doi: https://doi.org/10.1038/s41586-020-2405-7.

[6] Henrik Salje, Cécile Tran Kiem, Noémie Lefrancq, Noémie Courtejoie, Paolo Bosetti, Juliette Paireau, Alessio Andronico, Nathanaël Hozé, Jehanne Richet, Claire-Lise Dubost, Yann Le Strat, Justin Lessler, Daniel Levy Bruhl, Arnaud Fontanet, Lulla Opatowski, Pierre-Yves Boelle, and Simon Cauchemez. Estimating the burden of SARS-CoV-2 in France. Science, 369(6500):208–211, 2020.

[7] IHME COVID-19 Forecasting Team. Modeling COVID-19 scenarios for the United States. Nature Medicine, 27:94–105, 2021. doi: https://doi.org/10.1038/s41591-020-1132-9.

[8] Inga Holmdahl and Caroline Buckee. Wrong but useful—what COVID-19 epidemiologic models can and cannot tell us. New England Journal of Medicine, 383(4):303–305, 2020.

[9] Janice Hopkins Tanne. COVID-19: New York City deaths pass 1000 as Trump tells Americans to distance for 30 days. BMJ, 369:m1333, 2020. doi: https://doi.org/10.1136/bmj.m1333.

[10] Amit Uppal, David M Silvestri, Matthew Siegler, Shaw Natsui, Leon Boudourakis, R James Salway, Manish Parikh, Konstantinos Agoritsas, Hyung J Cho, Rajneesh Gulati, Milton Nunez, Anjali Hulbanni, Christine Flaherty, Laura Iavicoli, Natalia Cineas, Marc Kanter, Stuart Kessler, Karin V. Rhodes, Michael Bouton, and Eric K. Wei. Critical care and emergency department response at the epicenter of the COVID-19 pandemic. Health Affairs, 39(8):1443–1449, 2020. doi: https://doi.org/10.1377/hlthaff.2020.00901.

[11] Corinne N Thompson, Jennifer Baumgartner, Carolina Pichardo, Brian Toro, Lan Li, Robert Arciuolo, Pui Ying Chan, Judy Chen, Gretchen Culp, Alexander Davidson, et al. COVID-19 outbreak —-New York City, February 29–June 1, 2020. Morbidity and Mortality Weekly Report, 69(46):1725–1729, 2020.

[12] Qun Li, Xuhua Guan, Peng Wu, Xiaoye Wang, Lei Zhou, Yeqing Tong, Ruiqi Ren, Kathy S.M. Leung, Eric H.Y. Lau, Jessica Y. Wong, Xuesen Xing, Nijuan Xiang, Yang Wu, Chao Li, Qi Chen, Dan Li, Tian Liu, Jing Zhao, Man Liu, Wenxiao Tu, Chuding Chen, Lianmei Jin, Rui Yang, Qi Wang, Suhua Zhou, Rui Wang, Hui Liu, Yinbo Luo, Yuan Liu, Ge Shao, Huan Li, Zhongfa Tao, Yang Yang, Zhiqiang Deng, Boxi Liu, Zhitao Ma, Yanping Zhang, Guoqing Shi, Tommy T.Y. Lam, Joseph T. Wu, George F. Gao, Benjamin J. Cowling, Bo Yang, Gabriel M. Leung, and Zijian Feng. Early transmission dynamics in Wuhan, China, of novel coronavirus–infected pneumonia. New England Journal of Medicine, 382(13):1199–1207, 2020. doi: 10.1056/NEJMoa2001316.

[13] Joseph T Wu, Kathy Leung, and Gabriel M Leung. Nowcasting and forecasting the potential domestic and international spread of the 2019-nCoV outbreak originating in Wuhan, China: a modelling study. The Lancet, 395(10225):689–697, 2020. doi: https://doi.org/10.1016/S0140-6736(20)30260-9.

[14] Joseph T Wu, Kathy Leung, Mary Bushman, Nishant Kishore, Rene Niehus, Pablo M de Salazar, Benjamin J Cowling, Marc Lipsitch, and Gabriel M Leung. Estimating clinical severity of COVID-19 from the transmission dynamics in Wuhan, China. Nature Medicine, 26(4):506–510, 2020. doi: https://doi.org/10.1038/s41591-020-0822-7.

[15] Ruiyun Li, Sen Pei, Bin Chen, Yimeng Song, Tao Zhang, Wan Yang, and Jeffrey Shaman. Substantial undocumented infection facilitates the rapid dissemination of novel coronavirus (SARS-CoV2). Science, 368 (6490):489–493, 2020. doi: 10.1126/science.abb3221.

[16] Huaiyu Tian, Yonghong Liu, Yidan Li, Chieh-Hsi Wu, Bin Chen, Moritz U. G. Kraemer, Bingying Li, Jun Cai, Bo Xu, Qiqi Yang, Ben Wang, Peng Yang, Yujun Cui, Yimeng Song, Pai Zheng, Quanyi Wang, Ottar N. Bjornstad, Ruifu Yang, Bryan T. Grenfell, Oliver G. Pybus, and Christopher Dye. An investigation of transmission control measures during the first 50 days of the COVID-19 epidemic in China. Science, 368 (6491):638–642, 2020. doi: 10.1126/science.abb6105.

[17] Adam J Kucharski, Timothy W Russell, Charlie Diamond, Yang Liu, John Edmunds, Sebastian Funk, Rosalind M Eggo, and Centre for Mathematical Modelling of Infectious Diseases COVID-19 working group. Early dynamics of transmission and control of COVID-19: a mathematical modelling study. The Lancet Infectious Diseases, 20(5):553–558, 2020. doi: https://doi.org/10.1016/S1473-3099(20)30144-4.

[18] Kathy Leung, Joseph T Wu, Di Liu, and Gabriel M Leung. First-wave COVID-19 transmissibility and severity in China outside Hubei after control measures, and second-wave scenario planning: a modelling impact assessment. The Lancet, 395(10233):1382–1393, 2020. doi: https://doi.org/10.1016/S0140-6736(20)30746-7.

[19] Anthony Hauser, Michel J Counotte, Charles C Margossian, Garyfallos Konstantinoudis, Nicola Low, Christian L Althaus, and Julien Riou. Estimation of SARS-CoV-2 mortality during the early stages of an epidemic: A modeling study in Hubei, China, and six regions in Europe. PLoS Medicine, 17(7):e1003189, 2020. doi: https://doi.org/10.1371/journal.pmed.1003189.

[20] Jonas Dehning, Johannes Zierenberg, F Paul Spitzner, Michael Wibral, Joao Pinheiro Neto, Michael Wilczek, and Viola Priesemann. Inferring change points in the spread of COVID-19 reveals the effectiveness of interventions. Science, 369(6500):eabb9789, 2020. doi: 10.1126/science.abb9789.

[21] Kiesha Prem, Yang Liu, Timothy W Russell, Adam J Kucharski, Rosalind M Eggo, Nicholas Davies, Centre for the Mathematical Modelling of Infectious Diseases COVID-19 Working Group, Mark Jit, and Petra Klepac. The effect of control strategies to reduce social mixing on outcomes of the COVID-19 epidemic in Wuhan, China: a modelling study. The Lancet Public Health, 5:e261–e270, 2020. doi: https://doi.org/10.1016/S2468-2667(20)30073-6.

[22] H. Juliette T. Unwin, Swapnil Mishra, Valerie C. Bradley, Axel Gandy, Thomas A. Mellan, Helen Coupland, Jonathan Ish-Horowicz, Michaela A. C. Vollmer, Charles Whittaker, Sarah L. Filippi, Xiaoyue Xi, Meelodie Monod, Oliver Ratmann, Michael Hutchinson, Fabian Valka, Harrison Zhu, Iwona Hawryluk, Philip Milton, Kylie E. C. Ainslie, Marc Baguelin, Adhiratha Boonyasiri, Nick F. Brazeau, Lorenzo Cattarino, Zulma Cucunuba, Gina Cuomo-Dannenburg, Ilaria Dorigatti, Oliver D. Eales, Jeffrey W. Eaton, Sabine L. van Elsland, Richard G. FitzJohn, Katy A. M. Gaythorpe, William Green, Wes Hinsley, Benjamin Jeffrey, Edward Knock, Daniel J. Laydon, John Lees, Gemma Nedjati-Gilani, Pierre Nouvellet, Lucy Okell, Kris V. Parag, Igor Siveroni, Hayley A. Thompson, Patrick Walker, Caroline E. Walters, Oliver J. Watson, Lilith K. Whittles, Azra C. Ghani1, Neil M. Ferguson, Steven Riley, Christl A. Donnelly, Samir Bhatt, and Seth Flaxman. State-level tracking of COVID-19 in the United States. Nature Communications, 11(6189):1–9, 2020. doi: https://doi.org/10.1038/s41467-020-19652-6.

[23] Alberto Aleta, David Martin-Corral, Ana Pastore y Piontti, Marco Ajelli, Maria Litvinova, Matteo Chinazzi, Natalie E Dean, M Elizabeth Halloran, Ira M Longini Jr, Stefano Merler, Alex Pentland, Alessandro Vespignani, Esteban Moro, and Yamir Moreno. Modelling the impact of testing, contact tracing and household quarantine on second waves of COVID-19. Nature Human Behaviour, 4(9):964–971, 2020. doi: https://doi.org/10.1038/s41562-020-0931-9.

[24] Jonathan Fintzi, Damon Bayer, Isaac Goldstein, Keith Lumbard, Emily Ricotta, Sarah Warner, Lindsay M Busch, Jeffrey R Strich, Daniel S Chertow, Daniel M Parker, Bernadette Boden-Albala, Alissa Dratch, Richard Chhuon, Nichole Quick, Matthew Zahn, and Vladimir N Minin. Using multiple data streams to estimate and forecast SARS-CoV-2 transmission dynamics, with application to the virus spread in Orange County, California. arXiv preprint 2009.02654, 2020. URL https://arxiv.org/abs/2009.02654.

[25] Chloe Bracis, Eileen Burns, Mia Moore, David Swan, Daniel B Reeves, Joshua T Schiffer, and Dobromir Dimitrov. Widespread testing, case isolation and contact tracing may allow safe school reopening with continued moderate physical distancing: A modeling analysis of King County, WA data. Infectious Disease Modelling, 6:24–35, 2021.

[26] Thu Nguyen-Anh Tran, Nathan Wikle, Joseph Albert, Haider Inam, Emily R Strong, Karel Brinda, Scott M Leighow, Fuhan Yang, Sajid Hossain, Justin R Pritchard, Philip Chan, William P Hanage, Ephraim M Hanks, and Maciej F Boni. Optimal SARS-CoV-2 vaccine allocation using real-time seroprevalence estimates in Rhode Island and Massachusetts. medRxiv, 2021. doi: https://doi.org/10.1101/2021.01.12.21249694. URL https://www.medrxiv.org/content/10.1101/2021.01.12.21249694v1.

[27] Forrest W. Crawford, Zehang Richard Li, and Olga Morozova. COVID-19 projections for reopening Connecticut. medRxiv, 2020. doi: 10.1101/2020.06.16.20126425. URL https://www.medrxiv.org/content/early/2020/06/19/2020.06.16.20126425.

[28] Jordan Peccia, Alessandro Zulli, Doug E Brackney, Nathan D Grubaugh, Edward H Kaplan, Arnau Casanovas-Massana, Albert I Ko, Amyn A Malik, Dennis Wang, Mike Wang, Joshua L Warren, Daniel M Weinberger, Wyatt Arnold, and Saad B Omer. Measurement of SARS-CoV-2 RNA in wastewater tracks community infection dynamics. Nature Biotechnology, 38(10):1164–1167, 2020.

[29] State of Connecticut Office of Policy and Management. Connecticut Open Data: COVID-19 Data Resources. https://data.ct.gov/stories/s/COVID-19-data/wa3g-tfvc/. Accessed: 2021-03-29.

[30] Ned Lamont. Executive Order No. 7C: Protection of public health and safety during COVID-19 pandemic and response – further suspension or modification of statutes, March 2020. URL https://portal.ct.gov/-/media/Office-of-the-Governor/Executive-Orders/Lamont-Executive-Orders/Executive-Order-No-7C.pdf.

[31] Ned Lamont. Executive Order No. 7L: Protection of public health and safety during COVID-19 pandemic and response - extension of school cancellation, municipal retiree reemployment, open fishing season and additional public health measures, March 2020. URL https://portal.ct.gov/-/media/Office-of-the-Governor/Executive-Orders/Lamont-Executive-Orders/Executive-Order-No-7L.pdf.

[32] Ned Lamont. Executive Order No. 7X: Protection of public health and safety during COVID-19 pandemic and response – renter protections, extended class cancellation and other safety measures, educator certification, food trucks for truckers, April 2020. URL https://portal.ct.gov/-/media/Office-of-the-Governor/Executive-Orders/Lamont-Executive-Orders/Executive-Order-No-7X.pdf.

[33] Ned Lamont. Executive Order No 7II: Protection of public health and safety during COVID-19 pandemic and response – extension of school cancellation, home health care coverage, and food assistance measures, May 2020. URL https://portal.ct.gov/-/media/Office-of-the-Governor/Executive-Orders/Lamont-Executive-Orders/Executive-Order-No-7II.pdf.

[34] Ned Lamont. Executive Order No. 7H: Protection of public health and safety during COVID-19 pandemic and response - restrictions on workplaces for non-essential businesses, coordinated response effort, March 2020. URL https://portal.ct.gov/-/media/Office-of-the-Governor/Executive-Orders/Lamont-Executive-Orders/Executive-Order-No-7H.pdf.

[35] Centers for Disease Control and Prevention. COVID Data Tracker: Explore human mobility and COVID-19 transmission in your local area. https://covid.cdc.gov/covid-data-tracker/#mobility. Accessed: 2021-03-29.

[36] Ned Lamont. Governor Lamont releases rules for businesses under First Phase of Connecticut’s reopening plans amid COVID-19. State of Connecticut Press Release, May 9, 2020. URL https://portal.ct.gov/Office-of-the-Governor/News/Press-Releases/2020/05-2020/Governor-Lamont-Releases-Rules-for-Businesses-Under-First-Phase-of-Reopening-Plans.

[37] Ned Lamont. Governor Lamont releases business documents for Phase 2 reopening on June 17. State of Connecticut Press Release, June 7, 2020. URL https://portal.ct.gov/Office-of-the-Governor/News/Press-Releases/2020/06-2020/Governor-Lamont-Releases-Business-Documents-for-Phase-2-Reopening-on-June-17.

[38] Ned Lamont. Governor Lamont announces Connecticut moves toward Phase 3 reopening on October 8. State of Connecticut Press Release, September 24, 2020. URL https://portal.ct.gov/Office-of-the-Governor/News/Press-Releases/2020/09-2020/Governor-Lamont-Announces-Connecticut-Moves-Toward-Phase-3-Reopening-on-October-8.

[39] Ned Lamont. Governor Lamont provides update on Connecticut’s coronavirus response efforts. State of Connecticut Press Release, November 2, 2020. URL https://portal.ct.gov/Office-of-the-Governor/News/Press-Releases/2020/11-2020/Governor-Lamont-Coronavirus-Update-November-2.

[40] United States Census Bureau. American Community Survey (ACS), 2020. URL https://www.census.gov/programs-surveys/acs.

[41] Connecticut Hospital Association. URL https://cthosp.org/.

[42] Centers for Disease Control and Prevention. Scientific Brief: SARS-CoV-2 and Potential Airborne Transmission. https://www.cdc.gov/coronavirus/2019-ncov/more/scientific-brief-sars-cov-2.html, Updated Oct. 5, 2020.

[43] Forrest W. Crawford, Sydney Jones, Matthew Cartter, Samantha G. Dean, Joshua L. Warren, Zehang Richard Li, Jacqueline Barbieri, Jared Campbell, Patrick Kenney, Thomas Valleau, and Olga Morozova. Impact of close interpersonal contact on COVID-19 incidence: evidence from one year of mobile device data. medRxiv, 2021. doi: https://doi.org/10.1101/2021.03.10.21253282. URL https://www.medrxiv.org/content/10.1101/2021.03.10.21253282v1.

[44] Matt J Keeling and Pejman Rohani. Modeling infectious diseases in humans and animals. Princeton University Press, 2011.

[45] United States Census Bureau. 2010 cartographic boundary file, current block group for Connecticut. Data retrieved from http://magic.lib.uconn.edu/connecticut_data.html, 2010. Accessed: 2020-04-14.

[46] R Core Team. R: A Language and Environment for Statistical Computing. R Foundation for Statistical Computing, Vienna, Austria, 2020. URL https://www.R-project.org/.

[47] Karline Soetaert, Thomas Petzoldt, and R. Woodrow Setzer. Solving differential equations in R: Package deSolve. Journal of Statistical Software, 33(9):1–25, 2010.

[48] Centers for Disease Control and Prevention. COVID-19: When you’ve been fully vaccinated. https://www.cdc.gov/coronavirus/2019-ncov/vaccines/fully-vaccinated.html, Updated: 2021-04-02. Accessed: 2021-04-05.

[49] Robert Verity, Lucy C Okell, Ilaria Dorigatti, Peter Winskill, Charles Whittaker, Natsuko Imai, Gina Cuomo-Dannenburg, Hayley Thompson, Patrick GT Walker, Han Fu, Amy Dighe, Jamie T Griffin, Marc Baguelin, Sangeeta Bhatia, Anne Boonyasiri, Adhiratha andd Cori, Zulma Cucunubá, Rich FitzJohn, Katy Gaythorpe, Will Green, Arran Hamlet, Wes Hinsley, Daniel Laydon, Gemma Nedjati-Gilani, Steven Riley, Sabine van Elsland, Erik Volz, Haowei Wang, Yuanrong Wang, Xiaoyue Xi, Christl A Donnelly, Azra C Ghani, and Neil M Ferguson. Estimates of the severity of coronavirus disease 2019: a model-based analysis. The Lancet Infectious Diseases, 20(6):669–677, 2020. doi: https://doi.org/10.1016/S1473-3099(20)30243-7.

[50] Tegan K Boehmer, Jourdan DeVies, Elise Caruso, Katharina L van Santen, Shichao Tang, Carla L Black, Kathleen P Hartnett, Aaron Kite-Powell, Stephanie Dietz, Matthew Lozier, and Adi V Gundlapalli. Changing age distribution of the COVID-19 pandemic — United States, May–August 2020. Morbidity and Mortality Weekly Report, 69(39):1404–1409, 2020.

[51] Gabriel G Katul, Assaad Mrad, Sara Bonetti, Gabriele Manoli, and Anthony J Parolari. Global convergence of COVID-19 basic reproduction number and estimation from early-time SIR dynamics. PLoS ONE, 15(9): e0239800, 2020.

[52] Steven Sanche, Yen Ting Lin, Chonggang Xu, Ethan Romero-Severson, Nick Hengartner, and Ruian Ke. High contagiousness and rapid spread of severe acute respiratory syndrome coronavirus 2. Emerging Infectious Diseases, 26(7):1470–1477, 2020.

[53] Fiona P Havers, Carrie Reed, Travis Lim, Joel M Montgomery, John D Klena, Aron J Hall, Alicia M Fry, Deborah L Cannon, Cheng-Feng Chiang, Aridth Gibbons, Inna Krapiunaya, Maria Morales-Betoulle, Katherine Roguski, Mohammad Ata Ur Rasheed, Brandi Freeman, Sandra Lester, Lisa Mills, Darin S. Carroll, S. Michele Owen, Jeffrey A. Johnson, Vera Semenova, Carina Blackmore, Debra Blog, Shua J. Chai, Angela Dunn, Julie Hand, Seema Jain, Scott Lindquist, Ruth Lynfield, Scott Pritchard, Theresa Sokol, Lynn Sosa, George Turabelidze, Sharon M. Watkins, John Wiesman, Randall W. Williams, Stephanie Yendell, Jarad Schiffer, and Natalie J. Thornburg. Seroprevalence of antibodies to SARS-CoV-2 in 10 sites in the United States, March 23-May 12, 2020. JAMA Internal Medicine, 180(12):1576–1586, 2020.

[54] Shiwani Mahajan, Rajesh Srinivasan, Carrie A Redlich, Sara K Huston, Kelly M Anastasio, Lisa Cashman, Dorothy S Massey, Andrew Dugan, Dan Witters, Jenny Marlar, Shu-Xia Li, Zhenqiu Lin, Domonique Hodge, Manas Chattopadhyay, Mark D Adams, Charles Lee, Lokinendi V Rao, Chris Stewart, Karthik Kuppusamy, Albert I Ko, and Harlan M Krumholz. Seroprevalence of SARS-CoV-2-specific IgG antibodies among adults living in Connecticut: Post-infection prevalence (PIP) study. The American Journal of Medicine, In press, 2020. doi: https://doi.org/10.1016/j.amjmed.2020.09.024.

[55] Maciej F Boni. SARS-CoV-2 attack rates for Connecticut. https://twitter.com/maciekboni/status/1371847662077632513. Accessed: 2021-03-16.

[56] Shiwani Mahajan, César Caraballo, Shu-Xia Li, Yike Dong, Lian Chen, Sara K Huston, Rajesh Srinivasan, Carrie A Redlich, Albert I Ko, Jeremy S Faust, Howard P Forman, and Harlan M Krumholz. SARS-CoV-2 infection hospitalization rate and infection fatality rate among the non-congregate population in Connecticut. The American Journal of Medicine, In press, 2021. doi: https://doi.org/10.1016/j.amjmed.2021.01.020.

[57] Michelle A Waltenburg, Tristan Victoroff, Charles E Rose, Marilee Butterfield, Rachel H Jervis, Kristen M Fedak, Julie A Gabel, Amanda Feldpausch, Eileen M Dunne, Connie Austin, Farah S. Ahmed, Sheri Tubach, Charles Rhea, Anna Krueger, David A. Crum, Johanna Vostok, Michael J. Moore, George Turabelidze, Derry Stover, Matthew Donahue, Karen Edge, Bernadette Gutierrez, Kelly E. Kline, Nichole Martz, James C. Rajotte, Ernest Julian, Abdoulaye Diedhiou, Rachel Radcliffe, Joshua L. Clayton, Dustin Ortbahn, Jason Cummins, Bree Barbeau, Julia Murphy, Brandy Darby, Nicholas R. Graff, Tia K. H. Dostal, Ian W. Pray, Courtney Tillman, Michelle M. Dittrich, Gail Burns-Grant, Sooji Lee, Alisa Spieckerman, Kashif Iqbal, Sean M. Griffing, Alicia Lawson, Hugh M. Mainzer, Andreea E. Bealle, Erika Edding, Kathryn E. Arnold, Tomas Rodriguez, Sarah Merkle, Kristen Pettrone, Karen Schlanger, Kristin LaBar, Kate Hendricks, Arielle Lasry, Vikram Krishnasamy, Henry T. Walke, Dale A. Rose, Margaret A. Honein, and COVID-19 Response Team. Update: COVID-19 among workers in meat and poultry processing facilities —-United States, April–May 2020. Morbidity and Mortality Weekly Report, 69(27):887–892, 2020.

[58] Virginia E Pitzer, Melanie Chitwood, Joshua Havumaki, Nicolas A Menzies, Stephanie Perniciaro, Joshua L Warren, Daniel M Weinberger, and Ted Cohen. The impact of changes in diagnostic testing practices on estimates of COVID-19 transmission in the United States. MedRxiv, 2020. doi: https://doi.org/10.1101/2020.04.20.20073338. URL https://www.medrxiv.org/content/10.1101/2020.04.20.20073338v1.

[59] Iain Murray, Ryan Adams, and David MacKay. Elliptical slice sampling. In Proceedings of the thirteenth international conference on artificial intelligence and statistics, pages 541–548, 2010.

[60] Daniel P Oran and Eric J Topol. Prevalence of asymptomatic SARS-CoV-2 infection: a narrative review. Annals of Internal Medicine, 173(5):362–367, 2020.

[61] Diana Buitrago-Garcia, Dianne Egli-Gany, Michel J Counotte, Stefanie Hossmann, Hira Imeri, Aziz Mert Ipekci, Georgia Salanti, and Nicola Low. Occurrence and transmission potential of asymptomatic and presymptomatic SARS-CoV-2 infections: A living systematic review and meta-analysis. PLoS medicine, 17(9):e1003346, 2020.

[62] Centers for Disease Control and Prevention. COVID-19 Pandemic Planning Scenarios. https://www.cdc.gov/coronavirus/2019-ncov/hcp/planning-scenarios.html, 2021. Accessed: 2021-01-18.

[63] Hiroshi Nishiura, Tetsuro Kobayashi, Takeshi Miyama, Ayako Suzuki, Sung-Mok Jung, Katsuma Hayashi, Ryo Kinoshita, Yichi Yang, Baoyin Yuan, Andrei R Akhmetzhanov, and Natalie M Linton. Estimation of the asymptomatic ratio of novel coronavirus infections (COVID-19). International Journal of Infectious Diseases, 94:154–155, 2020.

[64] Kenji Mizumoto, Katsushi Kagaya, Alexander Zarebski, and Gerardo Chowell. Estimating the asymptomatic proportion of coronavirus disease 2019 (COVID-19) cases on board the Diamond Princess cruise ship, Yokohama, Japan, 2020. Eurosurveillance, 25(10):2000180, 2020.

[65] Timothy W Russell, Joel Hellewell, Christopher I Jarvis, Kevin Van Zandvoort, Sam Abbott, Ruwan Ratnayake, Stefan Flasche, Rosalind M Eggo, W John Edmunds, and Adam J Kucharski. Estimating the infection and case fatality ratio for coronavirus disease (COVID-19) using age-adjusted data from the outbreak on the Diamond Princess cruise ship, February 2020. Eurosurveillance, 25(12):2000256, 2020.

[66] Andrew William Byrne, David McEvoy, Aine B Collins, Kevin Hunt, Miriam Casey, Ann Barber, Francis Butler, John Griffin, Elizabeth A Lane, Conor McAloon, Kirsty O’Brien, Patrick Wall, Kieran A Walsh, and Simon J More. Inferred duration of infectious period of SARS-CoV-2: rapid scoping review and analysis of available evidence for asymptomatic and symptomatic COVID-19 cases. BMJ Open, 10(8), 2020. ISSN 2044-6055. doi: 10.1136/bmjopen-2020-039856. URL https://bmjopen.bmj.com/content/10/8/e039856.

[67] Wycliffe E Wei, Zongbin Li, Calvin J Chiew, Sarah E Yong, Matthias P Toh, and Vernon J Lee. Presymptomatic transmission of SARS-CoV-2 – Singapore, January 23–March 16, 2020. Morbidity and Mortality Weekly Report, 69(14):411, 2020.

[68] Matthew Biggerstaff, Benjamin J Cowling, Zulma M Cucunubá, Linh Dinh, Neil M Ferguson, Huizhi Gao, Verity Hill, Natsuko Imai, Michael A Johansson, Sarah Kada, Oliver Morgan, Ana Pastore y Piontti, Jonathan A. Polonsky, Pragati Venkata Prasad, Talia M. Quandelacy, Andrew Rambaut, Jordan W. Tappero, Katelijn A. Vandemaele, Alessandro Vespignani, K. Lane Warmbrod, Jessica Y. Wong, and the WHO COVID-19 Modelling Parameters Group. Early insights from statistical and mathematical modeling of key epidemiologic parameters of COVID-19. Emerging Infectious Diseases, 26(11), 2020. doi: https://doi.org/10.3201/eid2611.201074.

[69] Conor McAloon, Áine Collins, Kevin Hunt, Ann Barber, Andrew W Byrne, Francis Butler, Miriam Casey, John Griffin, Elizabeth Lane, David McEvoy, Patrick Wall, Martin Green, Luke O’Grady, and Simon J More. Incubation period of COVID-19: a rapid systematic review and meta-analysis of observational research. BMJ Open, 10(8):e039652, 2020. doi: 10.1136/bmjopen-2020-039652.

[70] Stephen A Lauer, Kyra H Grantz, Qifang Bi, Forrest K Jones, Qulu Zheng, Hannah R Meredith, Andrew S Azman, Nicholas G Reich, and Justin Lessler. The incubation period of coronavirus disease 2019 (COVID-19) from publicly reported confirmed cases: estimation and application. Annals of Internal Medicine, 172(9): 577–582, 2020.

[71] Jantien A Backer, Don Klinkenberg, and Jacco Wallinga. Incubation period of 2019 novel coronavirus (2019-nCoV) infections among travellers from Wuhan, China, 20–28 January 2020. Eurosurveillance, 25(5):2000062, 2020.

[72] Natalie M Linton, Tetsuro Kobayashi, Yichi Yang, Katsuma Hayashi, Andrei R Akhmetzhanov, Sung-mok Jung, Baoyin Yuan, Ryo Kinoshita, and Hiroshi Nishiura. Incubation period and other epidemiological characteristics of 2019 novel coronavirus infections with right truncation: a statistical analysis of publicly available case data. Journal of Clinical Medicine, 9(2):538, 2020.

[73] Jing Qin, Chong You, Qiushi Lin, Taojun Hu, Shicheng Yu, and Xiao-Hua Zhou. Estimation of incubation period distribution of COVID-19 using disease onset forward time: A novel cross-sectional and forward follow-up study. Science Advances, 6(33), 2020. doi: 10.1126/sciadv.abc1202. URL https://advances.sciencemag.org/content/6/33/eabc1202.

[74] Muge Cevik, Matthew Tate, Ollie Lloyd, Alberto Enrico Maraolo, Jenna Schafers, and Antonia Ho. SARS-CoV-2, SARS-CoV, and MERS-CoV viral load dynamics, duration of viral shedding, and infectiousness: a systematic review and meta-analysis. The Lancet Microbe, 2(1):e13–e22, 2021. doi: https://doi.org/10.1016/S2666-5247(20)30172-5.

[75] Kieran A Walsh, Karen Jordan, Barbara Clyne, Daniela Rohde, Linda Drummond, Paula Byrne, Susan Ahern, Paul G Carty, Kirsty K O’Brien, Eamon O’Murchu, Michelle O’Neill, Susan M Smith, Mairin Ryan, and Patricia Harrington. SARS-CoV-2 detection, viral load and infectivity over the course of an infection. Journal of Infection, 81(3):357–371, 2020. doi: https://doi.org/10.1016/j.jinf.2020.06.067.

[76] Xueting Qiu, Ali Ihsan Nergiz, Alberto Enrico Maraolo, Isaac I. Bogoch, Nicola Low, and Muge Cevik. The role of asymptomatic and pre-symptomatic infection in SARS-CoV-2 transmission — a living systematic review. Clinical Microbiology and Infection, 27(4):511–519, 2021. doi: https://doi.org/10.1016/j.cmi.2021.01.011.

[77] Rongrong Yang, Xien Gui, and Yong Xiong. Comparison of clinical characteristics of patients with asymptomatic vs symptomatic coronavirus disease 2019 in Wuhan, China. JAMA Network Open, 3(5):e2010182–e2010182, 2020.

[78] Kieran A Walsh, Susan Spillane, Laura Comber, Karen Cardwell, Patricia Harrington, Jeff Connell, Conor Teljeur, Natasha Broderick, Cillian F de Gascun, Susan M Smith, Mairin Ryan, and Michelle O’Neill. The duration of infectiousness of individuals infected with SARS-CoV-2. Journal of Infection, 81(6):847–856, 2020. doi: https://doi.org/10.1016/j.jinf.2020.10.009.

[79] Roman Wölfel, Victor M. Corman, Wolfgang Guggemos, Michael Seilmaier, Sabine Zange, Marcel A. Müller, Daniela Niemeyer, Terry C. Jones, Patrick Vollmar, Camilla Rothe, Michael Hoelscher, Tobias Bleicker, Sebastian Brünink, Julia Schneider, Rosina Ehmann, Katrin Zwirglmaier, Christian Drosten, and Clemens Wendtner. Virological assessment of hospitalized patients with COVID-2019. Nature, 581(7809):465–469, 2020.

[80] David Mc Evoy, Conor G McAloon, Aine B Collins, Kevin Hunt, Francis Butler, Andrew W Byrne, Miriam Casey, Ann Barber, John M Griffin, Elizabeth A Lane, Patrick Wall, and Simon J More. The relative infectiousness of asymptomatic SARS-CoV-2 infected persons compared with symptomatic individuals: A rapid scoping review. medRxiv, 2020. doi: https://doi.org/10.1101/2020.07.30.20165084. URL https://www.medrxiv.org/content/10.1101/2020.07.30.20165084v1.

[81] Daihai He, Shi Zhao, Qianying Lin, Zian Zhuang, Peihua Cao, Maggie H Wang, and Lin Yang. The relative transmissibility of asymptomatic COVID-19 infections among close contacts. International Journal of Infectious Diseases, 94:145–147, 2020.

[82] Centers for Disease Control and Prevention. Discontinuation of isolation for persons with COVID-19 not in healthcare settings. interim guidance. https://www.cdc.gov/coronavirus/2019-ncov/hcp/disposition-in-home-patients.html, 2020. Accessed: 2021-01-18.

[83] Shikha Garg, Lindsay Kim, Michael Whitaker, Alissa O’Halloran, Charisse Cummings, Rachel Holstein, Mila Prill, Shua J Chai, Pam D Kirley, Nisha B Alden, Breanna Kawasaki, Kimberly Yousey-Hindes, Linda Niccolai, Evan J Anderson, Kyle P Openo, Andrew Weigel, Maya L Monroe, Patricia Ryan, Justin Henderson, Sue Kim, Kathy Como-Sabetti, Ruth Lynfield, Daniel Sosin, Salina Torres, Alison Muse, Nancy M Bennett, Laurie Billing, Melissa Sutton, Nicole West, William Schaffner, H. Keipp Talbot, Aquino Clarissa, Andrea George, Alicia Budd, Lynnette Brammer, Gayle Langley, Aron J Hall, and Alicia Fry. Hospitalization rates and characteristics of patients hospitalized with laboratory-confirmed coronavirus disease 2019 – COVID-NET, 14 states, March 1–30, 2020. MMWR. Morbidity and Mortality Weekly Report, 69(15):458–464, 2020. doi: 10.15585/mmwr.mm6915e3.

[84] Pablo N Perez-Guzman, Anna Daunt, Sujit Mukherjee, Peter Crook, Roberta Forlano, Mara D Kont, Alessandra Løchen, Michaela Vollmer, Paul Middleton, Rebekah Judge, Chris Harlow, Anet Soubieres, Graham Cooke, Peter J White, Timothy B Hallett, Paul Aylin, Neil Ferguson, Katharina Hauck, Mark Thursz, and Shevanthi Nayagam. Clinical characteristics and predictors of outcomes of hospitalized patients with coronavirus disease 2019 in a multiethnic London national health service trust: a retrospective cohort study. Clinical Infectious Diseases, ciaa1091, 2020. doi: 10.1093/cid/ciaa1091.

[85] Nathan W Furukawa, John T Brooks, and Jeremy Sobel. Evidence supporting transmission of severe acute respiratory syndrome coronavirus 2 while presymptomatic or asymptomatic. Emerging Infectious Diseases, 26 (7), 2020.

[86] Jon C Emery, Timothy W Russel, Yang Liu, Joel Hellewell, Carl AB Pearson, CMMID COVID-19 Working Group, Gwen M Knight, Rosalind M Eggo, Adam J Kucharski, Sebastian Funk, Stefan Flasche, and Rein MGJ Houben. The contribution of asymptomatic SARS-CoV-2 infections to transmission on the Diamond Princess cruise ship. eLife, 9(e58699), 2020. doi: 10.7554/eLife.58699.

[87] Xi He, Eric H. Y. Lau, Peng Wu, Xilong Deng, Jian Wang, Xinxin Hao, Yiu Chung Lau, Jessica Y. Wong, Yujuan Guan, Xinghua Tan, Xiaoneng Mo, Yanqing Chen, Baolin Liao, Weilie Chen, Fengyu Hu, Qing Zhang, Mingqiu Zhong, Yanrong Wu, Lingzhai Zhao, Fuchun Zhang, Benjamin J. Cowling, Fang Li, and Gabriel M. Leung. Temporal dynamics in viral shedding and transmissibility of COVID-19. Nature Medicine, 26(5): 672–675, 2020.

[88] Mercedes Yanes-Lane, Nicholas Winters, Federica Fregonese, Mayara Bastos, Sara Perlman-Arrow, Jonathon R Campbell, and Dick Menzies. Proportion of asymptomatic infection among COVID-19 positive persons and their transmission potential: A systematic review and meta-analysis. PloS one, 15(11):e0241536, 2020.

[89] Marina Pollán, Beatriz Pérez-Gómez, Roberto Pastor-Barriuso, Jesús Oteo, Miguel A Hernán, Mayte Pérez-Olmeda, Jose L Sanmartín, Aurora Fernández-García, Israel Cruz, Nerea Fernández de Larrea, et al. Prevalence of SARS-CoV-2 in Spain (ENE-COVID): a nationwide, population-based seroepidemiological study. The Lancet, 396(10250):535–544, 2020.

[90] Helen Ward, Christina J Atchison, Matthew Whitaker, Kylie E. C. Ainslie, Joshua Elliott, Lucy C Okell, Rozlyn Redd, Deborah Ashby, Christl A. Donnelly, Wendy Barclay, Ara Darzi, Graham Cooke, Steven Riley, and Paul Elliott. Antibody prevalence for SARS-CoV-2 following the peak of the pandemic in England: REACT2 study in 100,000 adults. medRxiv, 2020. doi: 10.1101/2020.08.12.20173690. URL https://www.medrxiv.org/content/10.1101/2020.08.12.20173690v2.

[91] Barnaby Edward Young, Sean Wei Xiang Ong, Shirin Kalimuddin, Jenny G. Low, Seow Yen Tan, Jiashen Loh, Oon-Tek Ng, Kalisvar Marimuthu, Li Wei Ang, Tze Minn Mak, Sok Kiang Lau, Danielle E. Anderson, Kian Sing Chan, Thean Yen Tan, Tong Yong Ng, Lin Cui, Zubaidah Said, Lalitha Kurupatham, Mark I-Cheng Chen, Monica Chan, Shawn Vasoo, Lin-Fa Wang, Boon Huan Tan, Raymond Tzer Pin Lin, Vernon Jian Ming Lee, Yee-Sin Leo, David Chien Lye, and for the Singapore 2019 Novel Coronavirus Outbreak Research Team. Epidemiologic features and clinical course of patients infected with SARS-CoV-2 in Singapore. JAMA, 323 (15):1488–1494, 2020.

[92] The COVID-19 Investigation Team. Clinical and virologic characteristics of the first 12 patients with coronavirus disease 2019 (COVID-19) in the United States. Nature Medicine, 26(6):861–868, 2020. doi: https://doi.org/10.1038/s41591-020-0877-5.

[93] Enrico Lavezzo, Elisa Franchin, Constanze Ciavarella, Gina Cuomo-Dannenburg, Luisa Barzon, Claudia Del Vecchio, Lucia Rossi, Riccardo Manganelli, Arianna Loregian, Nicolò Navarin, Davide Abate, Manuela Sciro, Stefano Merigliano, Ettore De Canale, Maria Cristina Vanuzzo, Valeria Besutti, Francesca Saluzzo, Francesco Onelia, Monia Pacenti, Saverio G. Parisi, Giovanni Carretta, Daniele Donato, Luciano Flor, Silvia Cocchio, Giulia Masi, Alessandro Sperduti, Lorenzo Cattarino, Renato Salvador, Michele Nicoletti, Federico Caldart, Gioele Castelli, Eleonora Nieddu, Beatrice Labella, Ludovico Fava, Matteo Drigo, Katy A. M. Gaythorpe, Alessandra R. Brazzale, Stefano Toppo, Marta Trevisan, Vincenzo Baldo, Christl A. Donnelly, Neil M. Ferguson, Ilaria Dorigatti, Andrea Crisanti, and Imperial College COVID-19 Response Team. Suppression of a SARS-CoV-2 outbreak in the Italian municipality of Vo’. Nature, 584(7821):425–429, 2020.

[94] Seungjae Lee, Tark Kim, Eunjung Lee, Cheolgu Lee, Hojung Kim, Heejeong Rhee, Se Yoon Park, Hyo-Ju Son, Shinae Yu, Jung Wan Park, Eun Ju Choo, Suyeon Park, Mark Loeb, and Tae Hyong Kim. Clinical course and molecular viral shedding among asymptomatic and symptomatic patients with SARS-CoV-2 infection in a community treatment center in the Republic of Korea. JAMA Internal Medicine, 180(11):1447–1452, 2020.

[95] Muge Cevik, Krutika Kuppalli, Jason Kindrachuk, and Malik Peiris. Virology, transmission, and pathogenesis of SARS-CoV-2. BMJ, 371(m3862), 2020. doi: https://doi.org/10.1136/bmj.m3862.

[96] Liling Chaw, Wee Chian Koh, Sirajul Adli Jamaludin, Lin Naing, Mohammad Fathi Alikhan, and Justin Wong. Analysis of SARS-CoV-2 transmission in different settings, Brunei. Emerging infectious diseases, 26(11), 2020.

